# The temporal dynamics of the immune response to neoadjuvant androgen deprivation therapy suggests a window-of-opportunity for checkpoint inhibitor therapy in prostate cancer

**DOI:** 10.64898/2026.01.10.26343859

**Authors:** Anmbreen Jamroze, Renyuan Zhang, Kriti Ahuja, Lei Deng, Karan Jatwani, Uyen Nguyen, Bailey Farmer, Gaybrielle James, Michalis Mastri, Kevin H. Eng, Bo Xu, Yvonne M. Saenger, Yuanquan Yang, John J. Krolewski, Dean G. Tang, Gurkamal Chatta, Kent L. Nastiuk

## Abstract

**Purpose:** Novel therapies to prevent lethal castration resistant prostate cancer in response to standard-of-care androgen deprivation therapy (ADT) are required. Unfortunately, most prostate cancers are “immune cold” and fail to respond to checkpoint inhibitors (CPIs). To assess whether ADT induces changes that enable more effective CPI therapy, we examined the tumor immune micro-environment (TiME) following neoadjuvant ADT (nADT).

**Design:** Radical prostatectomy specimens from 43 nADT-treated patients were stratified into three duration groups and compared to each other and matched controls. RNA sequencing and quantitative multispectral immunofluorescence (qmIF) staining were performed to analyze transcriptomic and TiME abundance and cellular spatial relationship differences after nADT.

**Results:** Immune and inflammatory pathways, particularly of antigen presentation and adaptive immune response, were increased, most notably in tumors receiving 3-5 months nADT. qmIF revealed a complex temporal response in the TiME, with a dramatic influx of CTLs and T-helper cells after 3-5 months of nADT. However, after 6 months nADT, M2-like tumor associated macrophages (TAMs) and Tregs were strikingly increased while CTLs decreased. Spatially, CTLs and T-helper cells, clustered near tumor cells at 3-5 months nADT, were replaced by M2-TAMs in tumors receiving ≥ 6 months of nADT.

**Conclusion:** These data reveal the induction of a bi-phasic response in the TiME: robust CTL activation 3-5 months after nADT is initiated, followed by myeloid immunosuppression in tumors receiving prolonged nADT. This ADT-induced reprogramming of the TiME suggests a critical window of opportunity where short-duration ADT might augment CPI efficacy, converting cold into immunologically responsive tumors.

**Translational Relevance:** Immune Checkpoint inhibitors (CPIs) have not been effective in treating most human prostate cancers. This study describes the temporal dynamics of the immune response of primary prostate cancers to neoadjuvant androgen deprivation therapy (nADT), and suggests a strategic approach to improve the efficacy of CPIs in prostate cancer. After several months of nADT, inflammation and immune-related pathways were activated, accompanied by a robust infiltration of both CD8^+^ and CD4^+^ T cell into prostate tumors, indicating effector T cell education and activation. In contrast, six or more months nADT leads to an immunosuppressive shift, evidenced by increased M2-like tumor associated macrophages and regulatory T cells. Thus, our findings suggest a critical window of opportunity following nADT for initiating CPIs. This provides a rationale for the precise sequencing of nADT and CPI regimens to maximize therapeutic benefit.

## INTRODUCTION

The mainstay therapy for advanced and recurrent prostate cancer (PrCa) is androgen deprivation therapy (ADT). However, extended ADT leads to castration-resistant prostate cancer (CRPC), which has a poor prognosis. Trials of checkpoint inhibitor (CPI) therapies alone or in combination with androgen signaling pathway inhibitors in the CRPC setting, have not been effective (1–3). Indeed, fewer than 10% of CRPC tumors express an immune inflamed signature or other biomarkers predictive of CPI response, thus PrCa is classified as immune “cold” (4). However, the tumor immune microenvironment (TiME) in PrCa may be more aptly characterized as similar to the “immune excluded” immunophenotype described by Chan and Mellman (5). Indeed, we previously reported that cytotoxic T lymphocyte (CTL) density correlates with survival in primary PrCa (6), consistent with the possibility that immunosuppressed or partially-activated T-cells reside in primary prostate cancers.

Successful cancer immunotherapy depends on forming intratumoral immune triads of antigen-presenting cells, CD4^+^ helper and CD8^+^ T cells to educate T effector cells, enabling eradication of tumor (7). Androgens and other steroid hormones regulate the abundance and activity of most effector (T, NK, B) and regulatory (Treg, TAMs, MDSCs) immune cells, both normally and in disease (8). Because ADT is well-established as the primary therapy for advanced or recurrent PrCa, many investigation have sought to characterize the effects of ADT on the PrCa TiME. Multiple reports demonstrate that neoadjuvant ADT (nADT) induces a complex infiltrate containing each of these immune cell subtypes in PrCa (9–11), suggesting ADT might have immuno-activating effects. Additionally, androgens repress MHC expression, and hence ADT increases MHC-related genes, stimulating antigen presentation and anti-tumor immunity (12). However, this stimulation is transient, and MHC activity is subsequently repressed in CRPC (12). T cell activation occurs within a few weeks of nADT initiation in PrCa patients, as indicated by infiltration of CD4^+^ and CD8^+^ T cells, with increased effector cell response to tumor antigens (13, 14). Further, within two months of initiating nADT for locally advanced PrCa, immune and cytokine pathways, as well as CD8^+^ T cells, were enriched (15), highlighting the potential for nADT to alter immune modulation. In contrast, there have been multiple reports that ADT is accompanied by increasing Tregs and M2 macrophages (16–18), which correlate with poor prognosis for men with PrCa (11, 19). Overall, there are reports of both immune activation and suppression in response to ADT, but a lack of carefully executed temporal studies. Therefore, better defining ADT-induced changes in the prostate tumor microenvironment may enable combination therapy strategies to improve PrCa response to CPIs (20).

In this report, we analyzed transcriptomic profiles, imputed immune-cell content, and spatially identified immune-cell phenotypes and immune-cell temporal changes in prostatectomy specimens from a cohort of men treated with nADT prior to prostatectomy. Several months of nADT induced a transient immune activation that was followed by a compensatory immunosuppression. This suppressive state may render immune oncology therapeutics ineffective, and thereby drive PrCa resistance and recurrence. These findings illustrate the complex interplay between androgen signaling in the PrCa TiME and immune modulation. Importantly, nADT’s immune-activating properties suggest a limited-duration window of opportunity that might be a target for combination CPI therapy before an immunosuppressive TiME emerges.

## MATERIALS AND METHODS

### Ethics approval statement

The study was approved by the Institutional Review Board (IRB) at the Roswell Park Comprehensive Cancer Center (BDR 125520).

### Retrospective cohort

We identified patients with prostate cancer treated with ADT prior to radical prostatectomy performed at the Roswell Park Comprehensive Cancer Center. Many of these patients presented for a second opinion after initiating ADT (via an LHRH agonist) in the community. After a chart review to determine the duration of nADT exposure, clinical and pathologic data were abstracted from patient records. Data collected included age, race, year of prostatectomy, PSA levels, Gleason score, clinical and pathological stage. A cohort of 43 cases of men diagnosed with prostate cancer who received nADT for up to one year prior to prostatectomy was selected. These men underwent prostatectomy between 1995 and 2019 (mean 2006, interquartile range 6yr). Each patient had serum prostate-specific antigen (PSA) levels determined prior to initial nADT, prior to prostatectomy, and during long-term follow-up. A control cohort of 43 cases matched 1:1 on age, Gleason grade and stage, and year of prostatectomy who did not undergo nADT prior to prostatectomy was identified for comparative analysis (Table 1). Patients in either cohort who had a previous history of radiotherapy or chemotherapy were not accrued to this study.

**Table 1.** Clinical features and pathological data for cases/controls. P-values calculated using paired t-tests.

|  | nADT Cohort (43) | Controls (43) | P value |
| --- | --- | --- | --- |
| **Race** | % | % |  |
| White | 88.4 (38) | 90.7 (39) | 0.5698 |
| Black/AA | 11.6 (5) | 9.3 (4) |  |
| **Age** | Years | Years |  |
| Range of Ages | 50 - 71 | 48 - 69 | 0.4185 |
| Median Age at Surgery | 58 | 58 |  |
| **ADT** | % | % |  |
| 2 months or less | 41.9 (18) | 0 |  |
| 3 - 5 months | 32.5 (14) | 0 |  |
| 6+ months | 25.6 (11) | 0 |  |
| **PSA** | (ng/dL) | (ng/dL) |  |
| Pre-surgery PSA average | 9.8 | 7.1 | 0.1685 |
| Pre-surgery PSA range | 0.16 - 54.26 | 0 - 32.5 |  |
| **Clinical Staging** | % | % |  |
| Tx | 0 (0) | 2.3 (1) | 0.4977 |
| T1c | 62.8 (27) | 62.8 (27) |  |
| T2a-2c | 37.2 (16) | 32.6 (14) |  |
| T3b | 0 (0) | 2.3 (1) |  |
| **Pre-Surgery Gleason Score** | % | % |  |
| Gleason 6 | 25.6 (11) | 25.6 (11) | 0.2617 |
| Gleason 7 |  |  |  |
| 3+4=7 | 37.2 (16) | 30.2 (13) |  |
| 4+3=7 | 7.0 (3) | 14.0 (6) |  |
| Gleason 8 - 10 | 30.2 (13) | 30.2 (13) |  |
| **Pathological Staging** | % | % |  |
| pT1c | 0 (0) | 2.3 (1) | 0.05107 |
| pT2a-2c | 53.5 (23) | 72.1 (31) |  |
| pT3a | 27.9 (12) | 16.3 (7) |  |
| pT3a, N1 | 2.3 (1) | 0 (0) |  |
| pT3b | 11.6 (5) | 7.0 (3) |  |
| pT4 | 4.7 (2) | 2.3 (1) |  |
| **Post-Surgery Gleason Score** | % | % |  |
| Gleason 6 and under | 16.3 (7) | 13.9 (6) | 0.6497 |
| Gleason 7 |  |  |  |
| 3+4=7 | 30.2 (13) | 44.1 (19) |  |
| 4+3=7 | 25.6 (11) | 21.0 (9) |  |
| Gleason 8-10 | 25.6 (11) | 21.0 (9) |  |
| Unknown | 2.3 (1) | 0 (0) |  |

For each of the 86 cases, serial 5 µm thick sections were obtained from representative formalin-fixed, paraffin-embedded (FFPE) blocks and used for hematoxylin and eosin (H&E) staining, immunohistochemistry (IHC), and for quantitative multiplex immunofluorescence (qmIF) analysis, as well as for RNA and DNA extraction. H&E-stained slides were reviewed by a genitourinary pathologist to estimate tumor fraction, which ranged from ∼25%-75%. Transmembrane serine protease 2 (TMPRSS2) IHC staining (Clone AL20, cat. PT-mAb-AL20, 1:800 dilution) was performed on FFPE sections by the Roswell Park Pathology Network Shared Resource. Slides were digitally scanned and exported using Aperio ScanScope and ImageScope (Aperio Technologies, Inc.), and exported images analyzed for H-score using ImageJ software.

### Genomic mutational analysis

Whole transcriptome and targeted mutation analyses were performed using Agilent’s HS2 FFPE RNA reverse transcription and gDNA fragmentation and capture system using the Agilent SureSelect Cancer Comprehensive Genomic Profiling (CGP) Assay. The CGP panel targeted 679 genes for SNP analysis, 80 of which were also analyzed for gene fusions (including TMPRSS2), 26 for CNV (including BRCA1/2, MYC, and PTEN) and exons of 12 genes specifically for previously reported chromosomal translocations. Additionally, tumor mutational burden (TMB) and microsatellite instability (MSI) were measured for each sample. After capture, barcoded DNA was amplified, purified, and pooled. The pooled DNA was sequenced on an Illumina NovaSeq S1 flow cell using a standard paired-end 2x 150bp protocol. Sequencing was performed to an average depth of 40 million reads. Demultiplexed sequencing data of variants from each patient was analyzed for germline and somatic mutation using the Alissa Clinical Information platform (Agilent). SNVs were classified as likely pathogenic or not using ClinVar and a minimum 3% variant allele frequency was used to identify potentially significant variants. Since a paired peripheral germline comparison sample was not available for sequencing, 16 mutations per megabase DNA was used as the cut-off for TMB high, and 20% of markers showing instability as the cut-off for MSI high.

### RNA-sequencing and analysis

Total RNA from the 86 matched specimens was sequenced using Illumina’s NovaSeq6000. The quality of raw reads was assessed by FastQC v0.11.5 (http://www.bioinformatics.babraham.ac.uk/projects/fastqc/). Reads were mapped to the human reference genome GRCh38 (release 27) using STAR v2.6.0a (21). Gene expression was then quantified using RSEM (22). Estimated gene counts from RSEM were filtered and upper quartile normalized using edgeR (23). Differential gene expression analysis was performed using the limma package after voom transformation followed by linear regression (24). The significance threshold was set to false discovery rate (FDR) / adjusted p-value of less than 0.05 with a fold change greater than 2 (FDR < 0.05 and |log2 fold change| > 1). Transcripts per million (TPM) of selected genes were analyzed using R packages. The tumor immune microenvironment in the nADT and matched-control groups were compared using two independent tools, CIBERSORT (25) and ImmuCellAI (http://bioinfo.life.hust.edu.cn/web/ImmuCellAI/) (26). Gene set enrichment analysis (GSEA) (27) for the hallmark gene sets from the molecular signatures database (MSigDB) (28) was performed using the fgsea (29) Bioconductor package in R. GSEA for Gene Ontology (GO), Kyoto Encyclopedia of Genes and Genomes (KEGG), Wikipathways and Reactome Pathway gene sets was performed using the gseGO, gseKEGG, gseWP, and gsePathway functions of the R package clusterProfiler (30).

### Quantitative multiplex immunofluorescence analysis

Quantitative multiplex immunofluorescence staining was performed using the Vectra system (Perkin Elmer/Akoya Biosciences) to examine 5 µm sections of prostate tissue that were stained manually using the Opal 6-Plex Manual Detection Kit with tyramide signal amplification (Akoya Biosciences). The multiplex panel consisted of DAPI, CD3, CD8, CD68, HLA-DR, FoxP3, and NKX3.1 (detailed in Supplemental Table S1). The multiplex-stained slides were imaged using a Vectra 3 Polaris Automated Quantitative Pathology Imaging System. An initial quality control assessment of antibody expression in the qmIF panel was compared to that in single stained slides on control (tonsil) and prostate tissue by a pathologist. Staining patterns did not differ between the single- and multiplex scanned slides. This combination of markers distinguished cytotoxic T lymphocytes (CTLs), helper and regulatory T cells, tumor and stromal cells, and M1-and M2-like TAMs. Supplemental Table S2 defines the marker combinations used in this study.

The multispectral library slides were unmixed into eight channels using InForm software version 2.8: DAPI, HLA-DR-OPAL480, FOXP3-OPAL520, CD8-OPAL 570, CD68-OPAL 620, CD4-OPAL 690, NKX3.1-OPAL 780 and Auto Fluorescence and were fused with Phenoimager software version 3.0 to create a single q.tiff file for each sample. To reduce sampling-induced variability on tissue sections with infiltrative tumor, and to capture surrounding tumor-adjacent and normal prostate tissue TiME, regions-of-interest (ROIs) covering the whole tissue section were analyzed. After acquiring the panoramic low-magnification images at 4x, images encompassing the specimen using 1 mm^2^ sized regions-of-interest (ROIs) were acquired, and areas of poor quality and voids were excluded using Phenochart (PerkinElmer) software (31). In total, 1744 ROIs from 86 specimens were acquired for analysis. Cell phenotyping was based on biomarker expression within cells, nuclei and membranes using InForm Cell Analysis. Cells were then segmented into either tumor or stromal compartments, with NKX3.1 expression identifying tumor, using the inForm Tissue Finder tool. The density of each cell phenotype (tumor, stroma, and the delineated immune cell types) was then assessed (as cells/mm^2^).

### qmIF spatial analysis

The cellular neighborhood identification was performed using phenoptrReport (R, version 4.1.3), (https://akoyabio.github.io/phenoptr/). Specifically, image datasets, marked with specific cellular markers, and the “Cell Classification” tool were used to label and segment cell types. Supplemental Table S3 defines distances measured in this study. Distance between two cell subtypes was calculated using the x- and y-coordinates from the inForm raw data and nearest neighbor per-cell distance calculated using the Phenoptr GUI. A detailed spatial analysis was then conducted to examine the interactions between pairs of cell types (M2 to tumor and CTLs to M2, CTL to tumor, T-helper and CTL dyad to tumor (only if the CTL-T-helper cell dyad distance was less than 15 µm (7)) through a mixed radii nearest neighbor approach, capturing dynamic immune interactions and changes by treatment cohort. To identify shifts in immune cell relationships and their proximities over the course of treatment, the mixed radii nearest neighbor tool was used to measure neighboring cell proximities. Tabulated results showing the distribution and frequency of interactions at each radius across different treatment time points were generated using PhenoptrReports.

### Statistics and Bioinformatics

Clinical parameter and biomarker comparisons were carried out using paired t-tests in R. InForm statistical analyses of cell density changes between samples in treated and untreated cohorts were conducted using a paired T test using Origin Pro software (OriginLab Corp.). Correlation of CTL density (dichotomized as the upper quartile versus lower 75%-tile) with clinical outcome was determined by Kaplan-Meier testing with the alive patients censored, and a multivariate logistic regression model was used to examine the effect of CTLs density on patient survival.

### Data availability

In accordance with the Health Insurance Portability and Accountability Act deidentified data presented in this study are available upon request from the corresponding authors with institutional approval. However, deidentified exome and RNA-sequencing data are available in GEO. All R scripts used in this study are available from the authors upon request.

## RESULTS

### Clinical characteristics of cases and matched controls

Prostatectomy specimens from 86 men from 49 to 71 years old were included in this study; 43 of whom received neoadjuvant ADT prior to prostatectomy (Table 1). These 43 patients received a luteinizing hormone-releasing hormone (LHRH) antagonist, and 16 of these nADT treated patients also received the non-steroidal anti-androgen bicalutamide. As diagramed in Figure 1A, the nADT cohort was further divided into three subgroups based on treatment duration: Two months or shorter (n = 18), three to five months (n = 14), and six months or longer (n = 11). There were no differences in pre-treatment PSA levels between nADT duration groups (Supplemental Fig. S1A, and data not shown). Specimens from men who did not receive nADT, but were treated directly by prostatectomy were matched 1:1 with each of the nADT cases by age, race, stage/grade, and year of prostatectomy as a control cohort (n = 43). Both cohorts were comprised principally of white men (77/86). The majority of their prostate cancers (54/86) were stage T1c, and the men were younger than average for PrCa diagnosis (median age of 58 in both cohorts). Between the nADT and control cohorts, there were no differences in pre-treatment PSA, nor in demographics, biopsy Gleason grade or stage, nor were there any differences in the final stage or grade of the patient tumors recovered following prostatectomy (Table 1 and Supplemental Fig. S1A).

**Figure 1.**
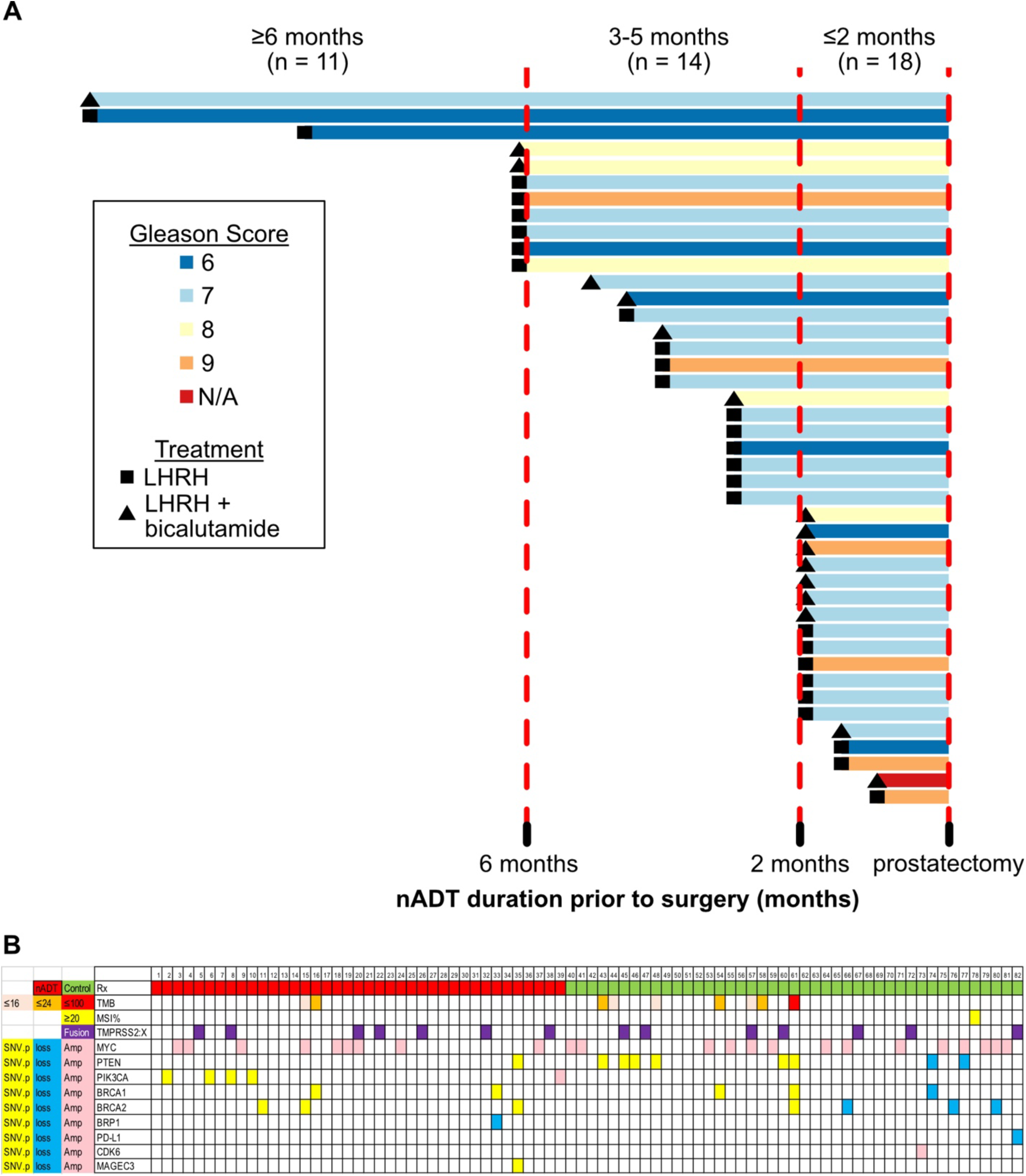
Duration of nADT exposure and genetic variant characterization of prostatectomy specimens from treated and matched control cohorts. **A.** Swimmer plot illustrating duration of nADT prior to prostatectomy. Specimen Gleason Score is indicated by colors, symbols indicate nADT therapy regimen, in the inset box. **B.** Table of samples indicating nADT exposure (n=39, red) or control samples (n=43, green), TMB quantitation (color indicates mutations per megabase, and MSI ≥ 20 (high). Pathogenic mutations (yellow), CNV loss/amplification (blue/pink), and gene fusions (purple) for the indicated genes, amongst 82 samples indicated across the top of the table.

### Time to biochemical recurrence, metastasis, and overall survival

The median PSA follow-up post-prostatectomy was 7.7 years (range 0.1 – 20.9) for the combined cohorts, 6.7 years for the nADT cohort, 9.7 years for the control cohort. Seven patients were lost to PSA follow up (two in the nADT cohort and five in the control cohort). Forty-one patients had biochemical recurrence (BCR), defined as PSA >0.2 ng/ml (20 in in the nADT cohort and 21 in the control cohort). The median time to BCR was 16.3 years in the nADT cohort, but was not reached in the control cohort (Supplemental Fig. S1B). Thirty-three patients (38.4%) developed metastases. The median time to metastasis (TTM) was 2.3 years (95% CI: 2.1 – 3.6), and did not differ between the cohorts. There were 10 deaths specifically from PrCa amongst these patients, five in the nADT cohort and three in the control cohort. The median overall survival (OS) was 16.1 years in the nADT cohort and 17.3 years in the untreated control cohort (Supplemental Fig. S1C).

### Genomic characteristics of nADT and control cohorts

Most (82/86) specimens from the nADT cases and controls yielded sufficient RNA and DNA for analysis of prostate cancer relevant genomic alterations. A total of 679 genes were assayed (see Materials and Methods) and 93% of samples yielded >100X coverage for these genes, with an effective genomic coverage of 1598 +/-2.3 Mb each. Clinical genomics analysis using the Agilent interpretive platform revealed frequent (18/82) MYC amplification, and a variety of lower frequency amplifications and deletions in cancer-related genes (Fig. 1B). The PTEN/PI3K pathway was also frequently affected (14/82), but this is likely an underestimate. Specifically, the CGP assay identified loss of PTEN in only two samples, suggesting technical limitations of the assay, as PTEN deletion has been reported to occur in up to 20% of prostate cancers (32). DNA repair genes (BRCA2, BRCA1, BRIP1) were disrupted in the germline of 5 patients, and somatically in an additional 7 samples. One sample displayed high MSI; another showed very high tumor mutational burden (TMB) and eight additional samples showed modest TMB (16-45 non-synonymous mutations or indels/MB). Terminating and frameshift mutations in PTEN, BRCA1 and BRCA2 were frequent in those samples with high TMB or MSI. TMPRSS2 fusions were identified in 14 samples (11 fusions with ERG, 1 each with ETV1, MRPS18C, and NBEAL1).

### TMPRSS2 expression was decreased by nADT

Since TMPRSS2 is androgen-regulated in prostate tumors (33, 34), we employed it as a biomarker to measure nADT efficacy. TMPRSS2 protein expression was quantitated by IHC staining of the 43 pairs of prostatectomy specimens from the nADT cohort and their matched controls (Supplemental Fig. 2A). Pair-wise comparison of the resulting H-scores indicates TMPRSS2 expression was reduced by nADT (Supplemental Fig. 2B). This reduction was sustained in the longer duration nADT groups (Supplemental Fig. 2C). Thus, TMPRSS2 protein expression indicates the nADT was effective in each of these duration groups prior to prostatectomy.

### nADT regulates AR, immune and inflammatory gene pathways in prostate tumors

To decipher the effects of nADT on the tumor, we then compared the transcriptomes from the 43 nADT samples to the paired control samples. Consistent with the IHC staining result, the RNA level of TMPRSS2 was strongly reduced and expression levels of other known AR-regulated genes, including KLK2, KLK3, KLK4 and AMACR were also decreased (blue in Fig. 2A). Expression of both chemokines CCL4/14/22 and chemokine receptors CCR4/7/10, important in T cell trafficking, were increased (red in Fig. 2A). The transcriptome data were then interrogated using GSEA to identify key molecular signaling or biological pathways altered by nADT. As expected, the ANDROGEN RESPONSE pathway was downregulated in nADT samples, which also showed enrichment of the INFLAMMATORY RESPONSE pathway (Fig. 2B). Further, Hallmarks of inflammation (TNFA SIGNALING VIA NFKB and IL2 STAT5 SIGNALING) were increased after nADT. To dissect immune-related signaling pathways activated by nADT, we used GSEA of the Gene Ontology (GO) gene sets to identify differences between the nADT-treated and the control tumor. As shown in Fig. 2C, nADT induced significant enrichment of immune effector GO pathways, particularly for antigen presentation and the adaptive immune response pathways. Additionally, regulation of lymphocyte and myeloid cell activation and migration GO gene sets were enriched by nADT (Supplemental Fig. S3A, S3B). Finally, GO analysis confirmed AR signaling was reduced in each of the duration groups of nADT treated tissues (Fig. 2D).

**Figure 2.**
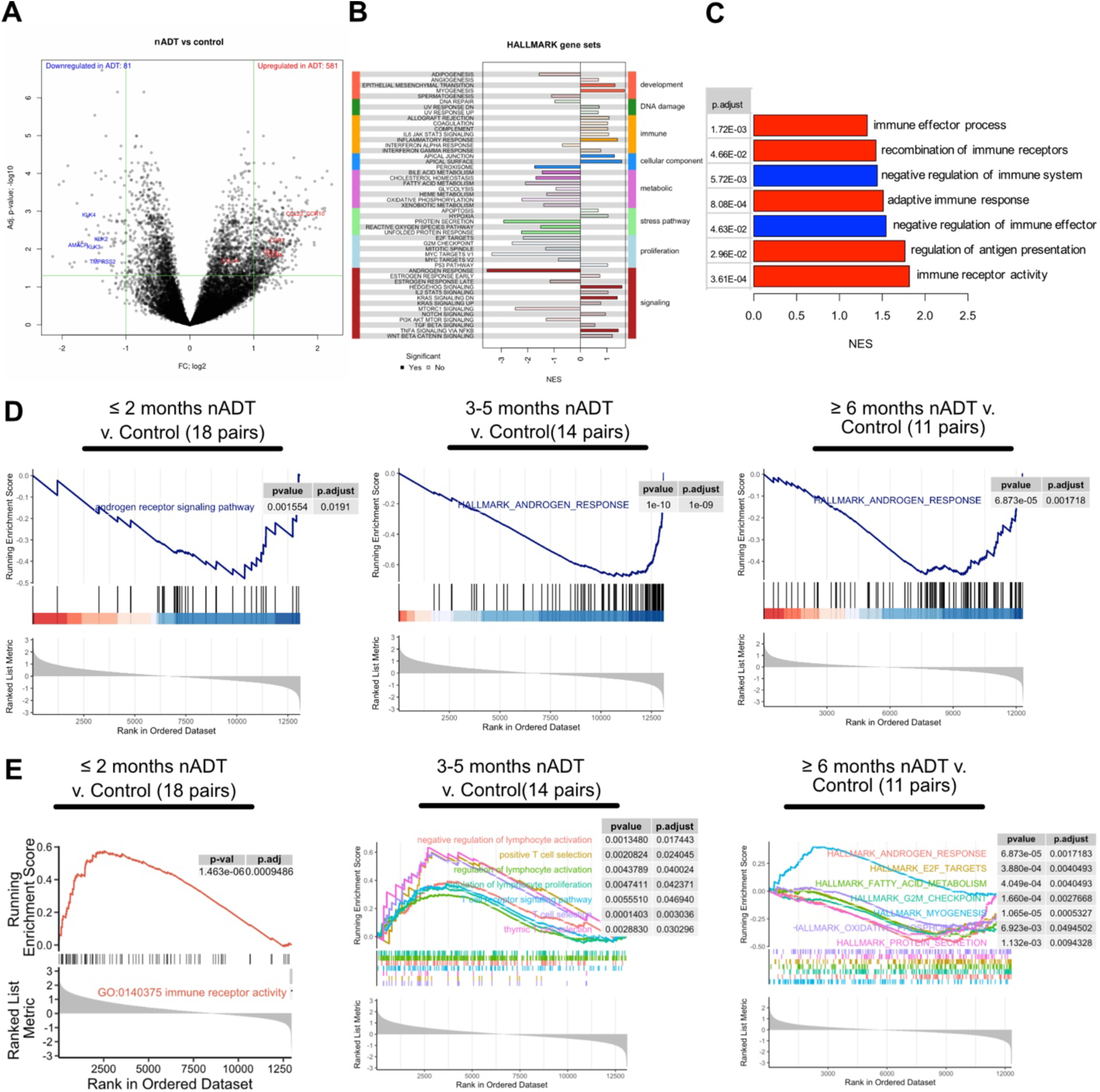
Differentially expressed genes and regulation of androgen and immune pathways in nADT cohort. **A.** Volcano plot of differentially expressed genes comparing nADT cohort to matched controls. Red indicates up-regulated and blue down-regulated genes. **B.** GSEA of hallmark gene sets for nADT cohort compared to matched controls. The deep colored (versus light colored) bars represents significantly enriched pathways. **C.** Enrichment of GO immune response pathways in nADT cohort compared to matched controls. Red indicates positive and blue negative enrichment of the indicated pathways. **D.** Enrichment of GO Androgen Receptor signaling pathway (GO,“GO:0030521) in the three nADT duration groups. **E.** Enriched GO and Hallmark pathways in the three nADT duration groups.

We next tested whether the immune response was duration dependent. GSEA Hallmark pathway analysis indicated enrichment of the INFLAMMATORY RESPONSE pathway in the ≤ 2 month nADT group and the INTERFERON GAMMA RESPONSE pathway in the 3-5 month nADT group, but these were reversed in the ≥ 6 month nADT group (Supplemental Fig. S3C-S3E). Interestingly, a comprehensive GSEA GO pathway analysis indicted immune related pathways, particularly lymphocyte activation, were enriched in both of the shorter duration nADT groups, while in the ≥ 6 month treated group, these were replaced by negative regulation of disparate growth-related pathways (Fig. 2E). Similarly, these same immune-related Hallmark pathways were most expressed in the 3-5 month nADT group pathways versus either the ≤ 2 month and ≥ 6 month nADT groups (Supplemental Fig. S3F, S3G).

### nADT alters the abundance of immune cell infiltrates in tumor microenvironment

To better characterize the functional outcome of nADT based on the altered immune-related pathways identified by the GO and Hallmark analysis, a paired analysis of the transcriptomes of the 43 nADT samples and their matched controls was performed to impute immune cell populations using the ImmuCellAI algorithm. Overall immune infiltration was increased by nADT, including an increase in antigen-presenting dendritic cells (Supplemental Fig. S4). As an alternative to the transcriptomic immune cell subset reconstruction algorithms, we evaluated the expression levels of marker genes characteristic of individual immune cells. Paired analysis, indicated the upregulation of CD8A, particularly in the ≤ 2 month nADT group (Fig. 3A, Supplemental Fig. S5A) and of the cytotoxic marker GZMB (Fig. 3B). CD4 expression was increased only in the 3-5 month nADT group (Fig. 3C, Supplemental Fig. S5C). The Treg marker FOXP3 expression was unchanged (Fig. 3D). In contrast, expression of genes encoding checkpoints PD-1 and TIM-3 were both increased, while those for TIGIT and LAG-3 were unchanged by nADT (Fig. 3E-3H). Expression of the macrophage marker CD68 was elevated after nADT (Fig. 3I). Expression of genes for markers of immune-suppressive M2-like macrophage CD163, CD204 (MSR1), and CD206 (MRC1) were each increased (Fig. 3J-3L). Expression of HLA-DRA, a marker of activated pro-tumorigenic M1 polarized macrophages, was overall unchanged, but increased specifically in the ≤2 month group and not with longer duration nADT (Fig. 3M, Supplemental Fig. S5M). These data suggest an initial immune activation and pro-inflammatory myeloid skewing, followed by an immunosuppressive tumor microenvironment after nADT driven by the M2-like myeloid population.

**Figure 3.**
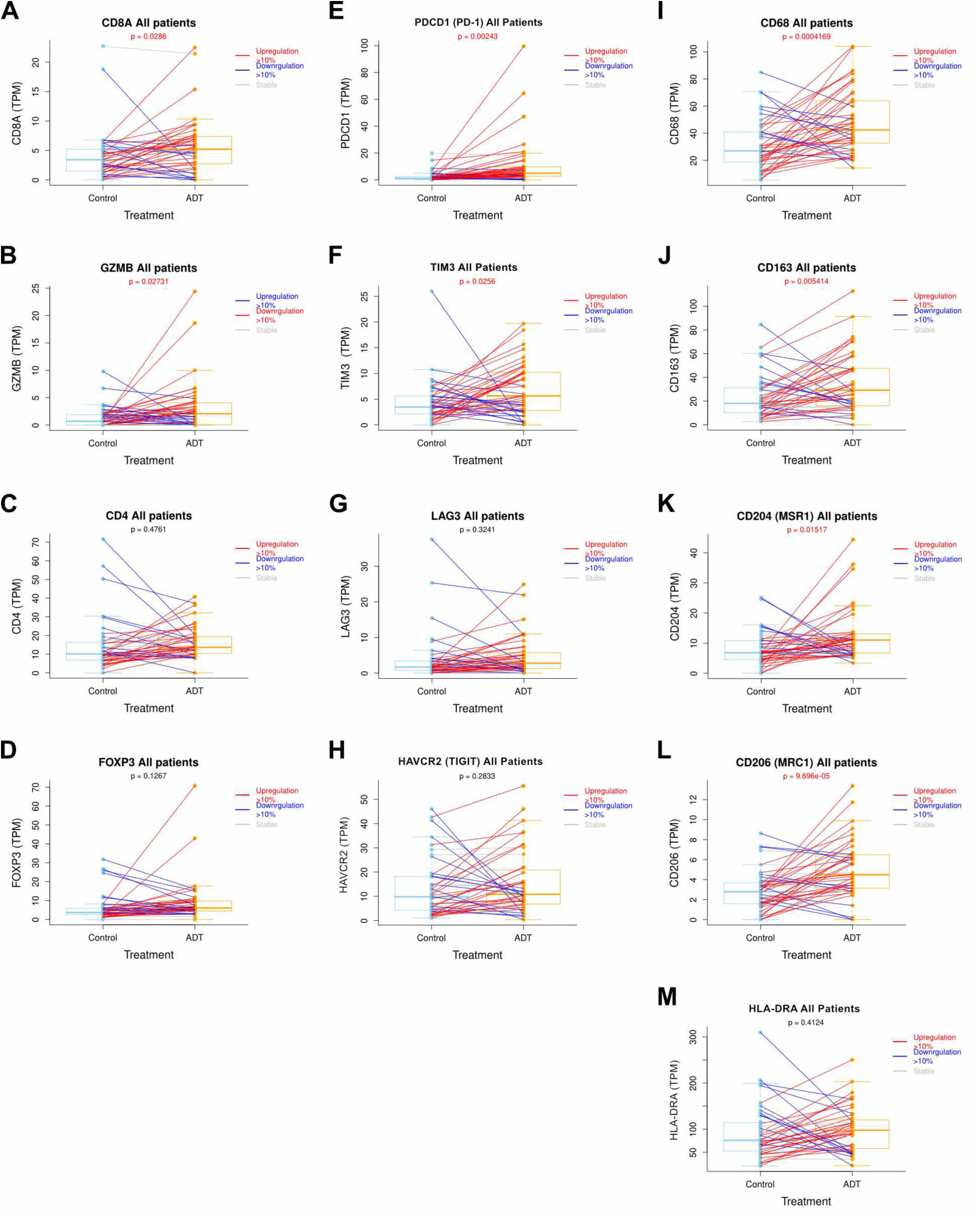
Immune marker gene expression after nADT. Comparison of transcripts per million (TPM) of the indicated genes in paired nADT (orange) and matched control tissues (blue) on left (43 pairs). Red lines connecting pairs indicate an increase in expression, while blue lines indicate a decrease after nADT. Paired t-test p-values are marked in red if < 0.05.

To test whether similar changes in the expression of mRNAs corresponding to TiME marker genes were observed with extended exposure to adjuvant ADT, we analyzed publicly available datasets of localized and advanced/metastatic CRPC (35–37). We found that following the long-term duration ADT that is a characteristic of CRPC treatment protocols, expression of the mRNAs for genes encoding immune checkpoints, including PD-1, LAG-3, TIGIT, and CD160, was increased (Supplemental Fig. S6A). Applying CIBERSORT analysis to these CRPC datasets, the increased expression of checkpoint genes in CRPC was accompanied by increases in CD8 T cells and Tregs, while estimates of memory and helper T-cell sub-sets, as well as activated NK cells, were decreased (Supplemental Fig. S6B). Expression of mRNAs encoding markers of suppressive (M2) macrophages, including IL-10, IL-13, VEGFA, and ARG1, were each increased in CRPC samples (Supplemental Fig. S6C). This was accompanied by an increase in the CIBERSORT estimate of M2 macrophages and a reciprocal decrease in M1 macrophages (Supplemental Fig. S6D). Finally, although the CIBERSORT estimate of CD8+ T cells increased in CRPC samples, the ratio of CD8+ T cells to M2 cells was decreased, whether using CIBERSORT immune cell population reconstruction, or measuring the ratio of mRNAs encoding CD8 and CD68 as specific markers of the corresponding cell populations (Supplemental Fig. S6E and S6F, respectively). Thus, overall we observed an increase in transcriptomic markers of immuno-suppressive cell populations in CRPC samples that received long-term ADT, similar to those in ≥ 6 month nADT treated specimens.

### Immune cell populations in the TME are altered by the duration of nADT

We then directly interrogated protein markers to determine the temporal and spatial patterns of immune cell populations in the TME in response to nADT. Specifically, we compared tissues from the nADT and control cohorts using Vectra qmIF staining to determine immune cell proportions and spatial interactions, as diagrammed in Supplemental Figure S7. We first examined the entire nADT (and control) cohorts (Supplemental Fig. S8). Cell type quantitation in prostatectomy specimens confirmed that nADT reduced tumor cell density while increasing stromal cell density consistent with cell death due to ADT. Interestingly, we observed a small but significant increase in cytotoxic T cell lymphocytes (CTLs) and Tregs in the nADT cohort, as well as M2-like TAMS, while M1-like TAMs were unchanged (Supplemental Fig. S8B).

Since we observed a mixed immune cell infiltrate of both CTLs and immunosuppressive myeloid cells when we examined the samples in the nADT cohort in aggregate, we next sought to examine the temporal effects of nADT duration on immune cell populations (Fig. 4). Visual inspection of representative immunofluorescent stained tissue sections (Fig. 4A) revealed that within 2 months of nADT, there was prominent M1-like TAMs (cyan) infiltration in the tumor stroma relative to matched control tissue, consistent with a pro-inflammatory response to tumor cell death. However, after 3-5 months nADT, CTLs (yellow) and T-helper cells (red) were dramatically more abundant, suggesting a profound immune activation. After 6 months nADT, there were few CTLs or T-helper cells, and a notable increase in both M2-like TAMs (green) and Tregs (white) around the tumor (magenta), suggesting a shift toward an immunosuppressive environment. These progressive changes in immune cell staining between each of the three duration of nADT groups, and from their matched controls (Fig. 4A) were validated by using image analysis software to quantitate all individual cells in the 1744 ROIs encompassing all of the prostate tissue in each of the 86 resection specimens (Fig. 4B). Notably, as observed visually, M1-like TAMs were increased in the ≤ 2 month nADT group, while CTLs and T-helper cells were broadly increased, particularly CTLs in tumors receiving 3-5 months nADT (Fig. 4B). The dynamic changes in cell proportions are summarized in Fig. 4C. However, after 6 months nADT, M2-like TAMs and Tregs were dramatically increased while CTLs were decreased (Fig. 4B-4C). Finally, the ratio of CTLs to M2-like TAMs and of CTLs to Tregs in nADT-treated and control samples were both decreased with longer nADT (Fig. 4D). Overall, these data suggest that nADT treatment initially leads to cell death-related inflammation, characterized by M1-like TAMS, followed by immune activation, with increased CD8+ T-cell infiltration, but that within six months nADT, the immune environment becomes suppressive, with increases in M2-like TAMs and Tregs, reflecting a shift from immune activation to suppression over the course of treatment.

**Fig. 4.**
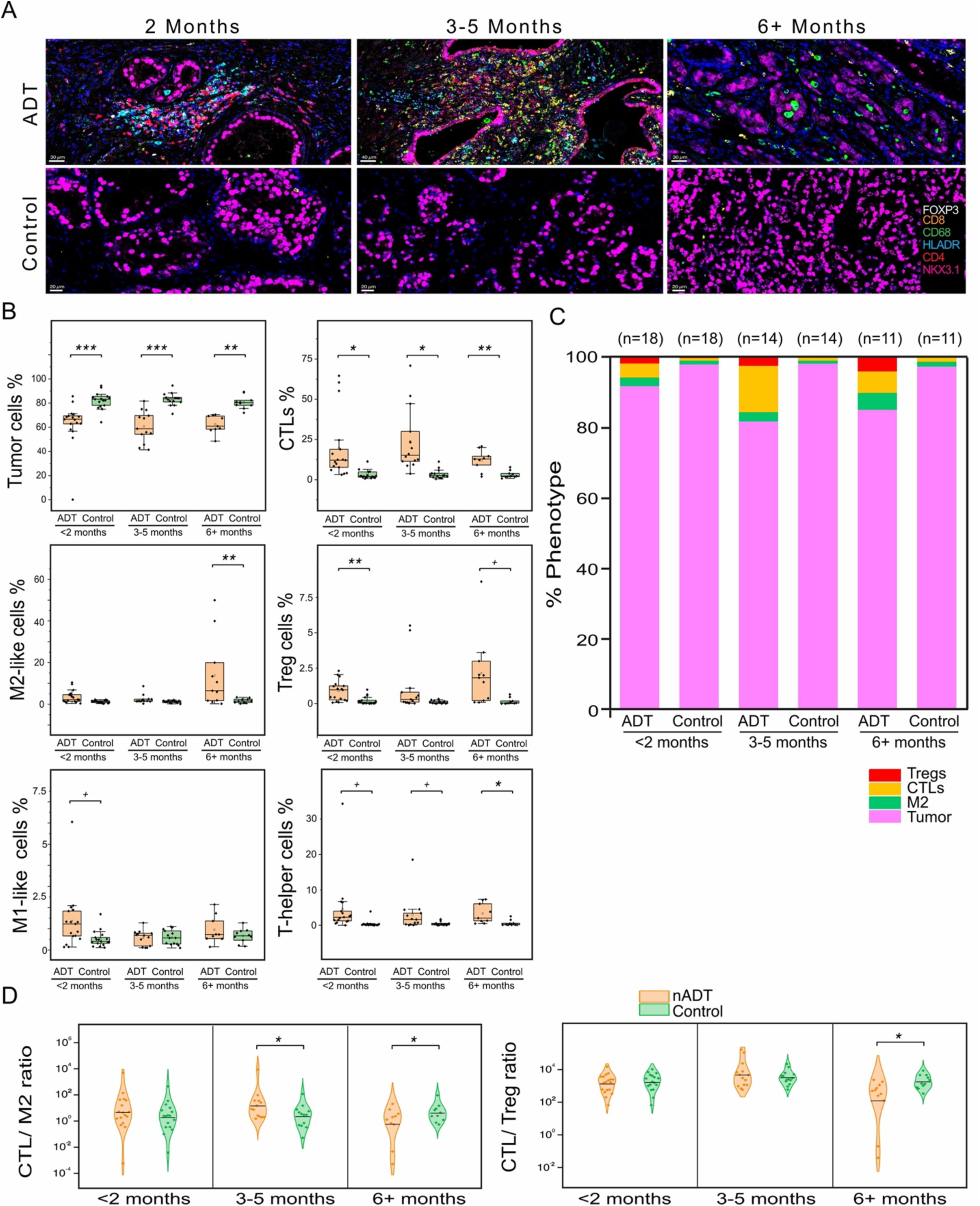
Extended nADT induced an increasingly suppressed TiME. **A.** Representative high power fields from 1744 qmIF images used to compare the distribution of immune cells in tumor between nADT-treated duration groups (≤ 2 months, 3-5 months, and ≥ 6 months) and matched control specimens. Marker identified by color indicated in inset, scale bar represents 20 µm. **B.** Bar charts quantitating changes in cell populations (as a percentage of total DAPI marked cells) for each duration nADT group (429, 356, 292 ROIs) vs matched control groups (322, 187, 158 ROIs). **C.** Stacked bars represent the proportion of each of the cell types (indicated in the color coded key below the stacked bar chart) for each nADT group and the matched control groups. **D.** Violin plots comparing the ratios of CTLs to M2-like TAMs (left) and CTLs to Tregs (right) in nADT-treated and control groups. Each data point represents the ratios from an individual patient sample, with comparisons made using the Wilcoxon signed-ranks test (+,*,**,*** indicates p-values of < 0.1, < 0.05; < 0.01; < 0.001, respectively).

### ADT induces immune cell populations to re-localized in the TME

Spatial analysis of the prostate TiME, identified by qmIF staining (Fig. 5), revealed dramatic changes in the distribution of CTLs that correlate with changes in T-helper and M2-like TAMs, as the duration of nADT increased. At ≤ 2 months nADT, M2-like TAMs were scattered or intraglandular, suggesting a phagocytic role and low immune suppressive activity. There was minimal immune cell infiltration, with CTLs and T-helper cells sparsely distributed and located far from the tumor, suggesting limited immune activation during the early phase of treatment. (Fig. 5A, left column). By 3-5 months nADT, there was a remarkable increase in both CTLs and T-helper cells (Fig. 5A, middle column), and these cells were found in close proximity to the tumor (Fig. 5A, middle column inset figures). This clustering resembles the immune cell triad of CD4+ and CD8+ T cells engaging with nearby antigen presenting cells, necessary for licensing cytotoxic T cells to mediate anti-tumor immunity (7). Later, at ≥ 6 months nADT, the immune landscape again shifted dramatically: there was a large reduction in the number of CD8+ T cells, and while CD4+ T cells remained clustered, fewer were proximal to the CTLs (Fig. 5A, third column). M2-like TAMs dramatically increased and infiltrated tumor glands (Fig. 5A, top row), and CTL interaction with tumor cells was dramatically reduced, suggesting a transition toward an immunosuppressive environment. Untreated matched control tissues showed no such changes (Supplemental Fig. S9A). While there was some heterogeneity in the predominant immunophenotype, ROIs from additional cases showed similar patterns of immune cells localization (Supplemental Fig. S10). More than 75% of the cases in the in the 3-5 month nADT group showed robust CTL staining, and more than 75% of the ≥ 6 month nADT group showed robust M2-like staining (Supplemental Table S4).

**Fig. 5.**
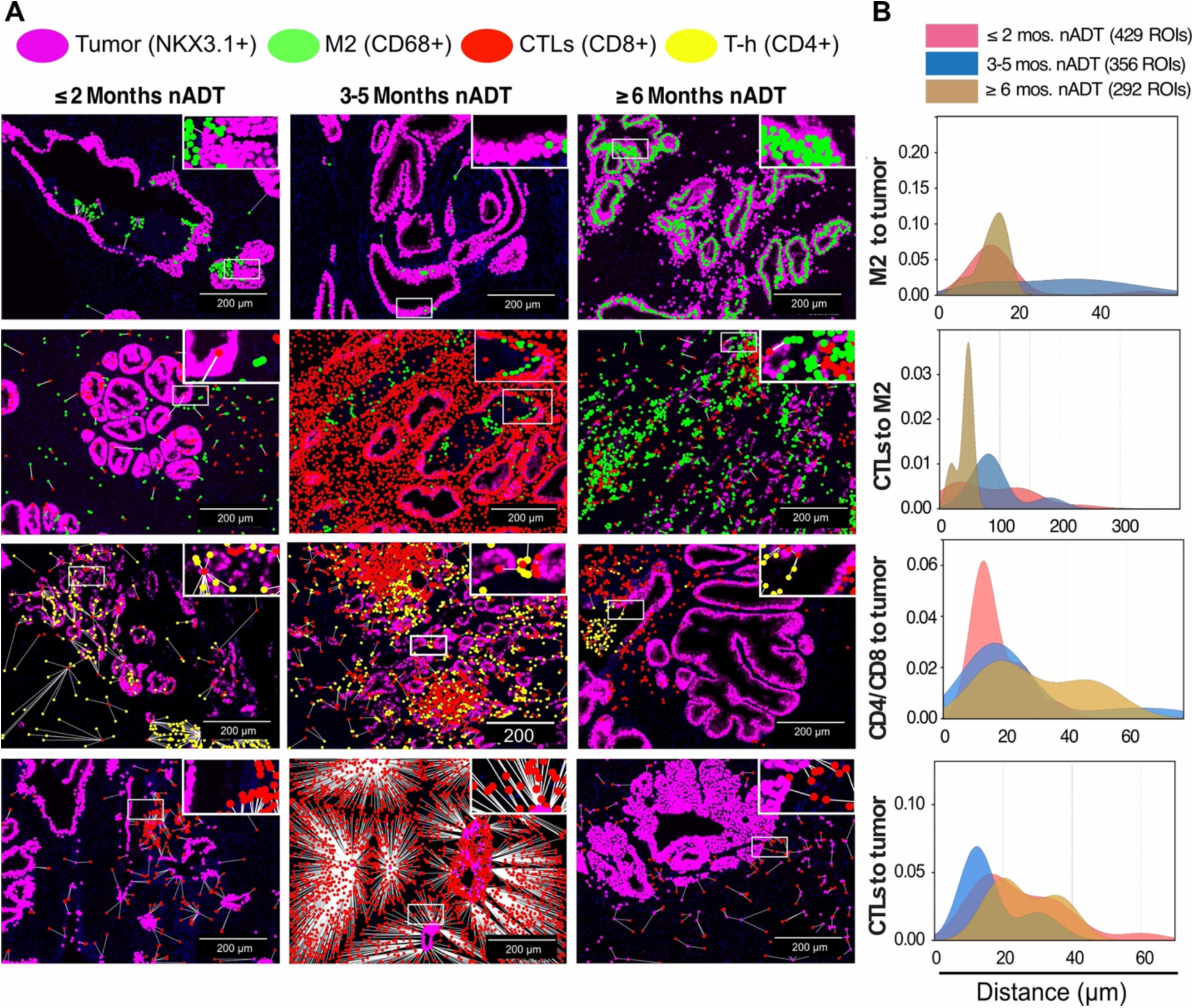
3-5 months nADT dispersed M2-like TAMs and increased T-helper and CTL proximity to tumor cells. **A**. Spatial mapping of immune cell distribution in nADT-treated prostatectomy specimens from each of the duration groups (columns). Example InForm cell mapping images from the samples nearest the mean of the distance distribution for each group illustrating localization of CD68^+^, HLA-DR^-^ M2-like TAMs and their distance to NK3.1^+^ tumor (top row); CD8^+^ CTLs and the distance to nearest M2-like cell (second row); CD4^+^/CD8^+^ immune cell dyad to nearest tumor cell (third row); CD8^+^ CTLs and the distance to nearest tumor cell (bottom row). **B.** Density plots showing temporal changes in cell-cell proximities in each of the ROIs from the indicated (by color) groups of nADT-treated samples. Matched control examples and distances for each nADT group in Supplemental Figure S9. Additional nADT and control examples in Supplemental Figure S10.

Quantitation of the spatial distribution of these cells in the tumor tissues revealed dynamic changes with nADT. The nearest neighboring cell-cell interaction distances, visually depicted by white lines in Fig. 5A, were quantitated in density plots of cell proximities in the 1077 ROIs from the nADT cohort resection specimens (Fig. 5B). M2-like TAM proximity to tumor cells was variable based on duration of nADT. In the ≤ 2 month nADT group, M2-like TAMs were largely intraglandular and close to tumor cells, (maxima 13 µm, pink peak in Fig. 5B top panel). In contrast, M2-like TAM proximity to tumor cells showed a wide distribution by 3-5 months nADT (centered around 35 µm, blue in Fig. 5B top panel), suggesting reduced immunosuppressive influence. However, after ≤ 6 months nADT, M2-like TAMs were again proximal to tumor cells (brown peak at 15 µm in Fig. 5B top panel). The distance between CTLs and M2-like TAMs in specimens from the ≤ 2 month nADT group was widely distributed, but by 3-5 months nADT, this separation was further increased (by ∼50 µm), suggesting reduced interaction and enhanced CTL activity (Fig. 5B, pink and blue, second panel). Similar to the M2 to tumor distance, after 6 months nADT, the CTL to M2 distance dramatically decreased (20-50 µm, Fig. 5B, brown, second panel), suggesting regions of continued immune suppression and CTL exhaustion. The clustering of dyads of T helper cells and CTLs with adjacent tumor cells is a proxy for the activated anti-tumor immune triads (7). These T-helper/CTL dyads were closely associated (≤ 20 µm) with adjacent tumor cells in specimens from both the ≤ 2 month nADT and 3-5 month nADT groups, suggesting coordinated immune activation (Fig. 5B, pink and blue curves, third panel). However, by ≥ 6 months nADT, the proximity decreased, with bimodal peaks at 20 µm, and a new peak at 50 µm (Fig. 5B, third panel, brown curve). This progression from tumor-immune interaction dense cell subsets early in the course of nADT to the increased proximity of suppressive M2-like TAMs to tumor cells later is apparent visually (Fig. 5A, bottom panels) and is quantitatively reflected in the large number of closely interacting CTLs in the TiME of the 3-5 month nADT group, which is greatly reduced by ≥ 6 months nADT (Fig. 5B, bottom panel). These alterations in nearest neighbor proximities in the nADT specimens were not evident in ROIs from the matched control tissues (Supplemental Fig. S9). Importantly, the significance of each of the proximity findings in Figure 5B were confirmed by spatial distribution analysis of all 1744 ROIs representing the 43 individual nADT and 43 individual matched control patient resections specimens (Supplemental Fig. S9B).

## DISCUSSION

While current guidelines do not support nADT prior to radical prostatectomy, the tissue archives at Roswell Park Comprehensive Cancer Center accumulated prostatectomy specimens from patients treated with neoadjuvantally with ADT for 1-12 months (Fig. 1). Endpoint transcriptomic and spatial quantitative immunofluorescent profiling of these specimens provided an opportunity to investigate the temporal evolution of the PrCa TiME, as might occur in advanced, recurrent, or metastatic PrCa. Validation of nADT efficacy was determined using TMPRSS2 expression as a biomarker of ADT effect in the treated cohort, and found to be reduced for each of the duration groups (Supplemental Fig. S2). nADT was previously reported to downregulate TMPRSS2 mRNA expression in resections after seven days (38) and one month (39). To our knowledge, this is the first demonstration of parallel reduction of both TMPRSS2 mRNA and protein, and of a reduction over longer durations of nADT in patient specimens, suggesting wide utility as a biomarker of ADT. Overall, nADT induced a loss of tumor cells with a corresponding increase in the proportion of stroma, accompanied by increased immune infiltrate (Supplemental Fig. S8).

In addition to effective AR suppression, transcriptomic pathway analysis revealed inflammatory pathway response (Fig. 2B-2E). Quantitation of the immune cells in the TiME revealed an increase in M1-like TAMs in the ≤ 2 month nADT group that accompanied a decrease in tumor cells (Fig. 4B). These observations are consistent with tumor cell death, unequivocally the clinical goal of ADT. The early increase in M1-like TAMs is also consistent with an immunogenic cell death (ICD) model for the earliest immune response to ADT. ICD, induced by pro-apoptotic anti-cancer therapies, is a type of tumor cell death that elicits an anticancer immune response relying on the activation of damage-associated molecular patterns (DAMPs) (40). Antigens from dying cancer cells promote the recruitment and activation of antigen-presenting cells (APCs), especially dendritic cells (DCs) and M1-type TAMs. DAMPs consequently enhance DC maturation and DCs ability to stimulate transient immune effector T cells (41).

This initial period of ICD in response to nADT is followed by a window of intense immune activation. This was confirmed by early (≤ 2 months) upregulation of cytotoxic T cell markers (e.g., CD8A, GZMB; Fig. 3), followed by a dramatic increase in CTLs by 3-5 months nADT (Fig. 4). This correlated with GSEA evidence of increased T-cell trafficking and IFNG/TNF signaling to sustain T cell activation (Fig. 2E), and increased dendritic cells which may mediate antigen presentation (Supplemental Fig. S4B) following 3-5 months nADT. Similar transcriptomic changes in Hallmark gene sets of androgen signaling, as well as increases in IFNG and TNF signaling have been reported after enzalutamide therapy (42, 43). The early active immune response was confirmed by spatial qmIF analysis which revealed proximal paired T-helper cells and CTLs near tumor cells (Fig. 5A, left column). When combined with antigen presenting dendritic cells (Fig. S4B), this cell triad clustering might mediate active licensing of CD8^+^ T cells to effect anti-tumor immunity (7). These T-helper/CTL dyads moved closer to tumor by 3-5 months nADT, when an immune “storm” of CD8^+^ T cells adjacent to both T helper cells and tumor was apparent (Fig. 5A middle column).

However, after ≥ 6 months nADT, the PrCa TiME becomes immunosuppressive. Both Tregs and M2-like TAMs were increased (Fig. 4B). The overall proportion of CTLs was reduced, as were the ratios of CTLs to M2-like TAMs and CTLs to reg cells (Fig. 4D). The CTLs were juxtaposed to the M2-like TAMs, but more distant from tumor cells (Fig. 5). These M2-like TAMs are anti-inflammatory and pro-tumorigenic (44), and predict poor survival post-ADT (45). In the specimens from the ≥ 6 month nADT group, the M2-like TAMs appear to form a protective layer around the tumor glands (Fig. 5A). These observations in the ≥ 6 month nADT group are consistent with reports that on progression to CRPC, the TiME contains high levels of interferon and activated but dysfunctional CD8^+^ T cells, and the tumors are resistant to CPIs (2, 46).

Several other reports demonstrate ADT induces T cell activation in PrCa tumors. ADT (or radiation) results in autoantibodies in 25%-30% patients (13). During the first month after nADT prior to resection, markers of CD4+ and CD8+ T cells increased, with peak CD3+ T cell activity at three weeks (10). Within one month of nADT, circulating naïve T cells increase, with an increase in effector cell response (14). Similar transient activation of immune response has also been observed in mouse models (10). Combining a GM-CSF-secreting vaccine with nADT increased CD8+ T-cell infiltration, but also raised levels of Tregs (16). This finding underscored the need for strategies targeting Tregs to improve therapeutic efficacy (20). AR signaling activity suppresses hallmark interferon response and suppresses T cell activity, and ADT enhances response to checkpoint blockade therapy in mouse melanoma and prostate cancer models (46), analogous to the interferon associated checkpoint responsiveness in human colorectal cancer (47). Since ADT reduces immune tolerance to prostate-expressed antigens in mice, allowing T cells to develop effector function in response to vaccination (48), our data suggests that ADT might best be employed several months prior to immunotherapy with CPIs.

Independent of CPIs, the immune activation and subsequent suppression was consequential in this nADT cohort. CTL density was prognostic for OS in patients treated with fewer than six months of nADT, however the effect was lost with the longer duration nADT (Supplemental Fig. S11), in accord with the above temporal characterization. This is in agreement with our previous finding of a correlation of CTL density with PFS and OS in much larger (n=230) cohort of primary resection specimens (6). Indeed, the finding is in accord with a report that the combination of CTL and PD-L1 expression predicts biochemical recurrence (49). Thus, the density of immune cells and their proximity to tumor cells may serve as an independent prognostic biomarkers of clinical outcomes.

Limitations of the study include the nADT cohort sample size, particularly of the duration sub-groups, the retrospective providential nature of the cohort, and extended period of time required to gather sufficient samples for analysis. Many of the patients presented at a comprehensive cancer center for a second opinion in the mid-2000s, having been previously treated in the community with ADT. These men may have received neoadjuvant therapy in response to perceived risk at presentation. However, there was no difference between the groups in PSA levels determined pre-ADT, or in survival (Supplemental Fig. S1). Further, there was no substantive difference in the genetic profile between the nADT treated cohort and the untreated control cohort (Fig. 1B). To control for any changes due to evolving practice, each nADT sample was paired with a control sample of identical stage/grade and year of resection. The few patients with higher TMB and MSI are not amongst the examples presented in Figure 5, and were more prevalent in the untreated control cohort (Fig. 1B). Finally, qmIF analysis using the Vectra system restricted the panel of antibodies to six. This necessarily limited the depth of the TiME cell phenotyping, and future studies might employ more extensive panels to, for example, directly characterize the extent of T cell exhaustion.

In conclusion, long-term ADT remodels the TiME in PrCa with a transient immune activation and then immune suppression which leads to the occurrence of CRPC that is also CPI-resistant (Supplemental Fig. S12A). One approach to prevent the development of CRPC is to improve the initial treatment of patients with advanced PrCa by identifying windows of opportunity to combine CPIs with ADT. The present examination of nADT duration groups enabled dissection of the time-course of changes in the TiME (Supplemental Fig. S12B). These findings align with an ICD model, with DAMPs released from dying tumor cells activating APCs and M1 macrophages, boosting T cell responses, enhancing anti-tumor immunity by promoting CTL priming and sustained immune engagement, supporting the early robust immune activation observed in our study. We hypothesize that in response to the initial immune activation phase, there is a reciprocal induction of an immune suppression phase (Supplemental Fig. S12, blue line) marked by checkpoint expression, Treg and myeloid cell recruitment resulting in effector T cell suppression (Supplemental Fig. S12C). Reprogramming the TiME by modifying production of chemokines and cytokines is an emerging strategy to convert cold tumors into immunologically responsive tumors and improve patient outcomes. It is not just the quality and the quantity of CTLs, but also the spatial positioning of immune cells in tumors that is critical for long-term tumor control (7). Indeed, we found dramatic changes in the distribution of CTLs over time of exposure to nADT, suggesting large differences in potential response to immune-directed therapies early or late in the course of ADT.

Our findings provide crucial insights into the temporal dynamics of ADT-induced immune modulation in PrCa. By identifying a period of high T cell infiltration after several months of nADT but a subsequent increase in immunosuppressive Tregs and M2 TAMs after six months nADT, we highlight a critical window for therapeutic intervention. In advanced and recurrent patients initiating ADT, delivering CPIs in this early phase of ADT-induced immune activation may optimize treatment efficacy. Moreover, understanding the shift toward immune suppression over time opens avenues to counteract M2 or Treg mediated immune suppression that could be sequenced with longer duration ADT (50–52), potentially improving long-term outcomes in prostate cancer management.

## Acknowledgements

We thank the patients whose specimens and data made this research possible; Felicity Hall, Mike Ruvolo, and Brigette Kipphut at Agilent for early access to the CGP assay and assistance with data interpretation; Prashant Singh and Philip Galbo at the Roswell Park Genomics shared resource for assistance with sample preparation and sequencing; Anges Witkiewicz and the Advanced tissue imaging shared resource, and the staff of the Roswell Park Pathology and Data shared resources.

## Supplemental Tables

**Supplemental Table S1.** Antibodies used in quantitative multispectral immunofluorescent analysis.

|  | Antigen | Supplier | Dilution | Clone | Cat# | Color (em. nm) |
| --- | --- | --- | --- | --- | --- | --- |
| 1 | **DAPI** |  |  |  |  | Blue (460) |
| 2 | **CD4** | Abcam | 1:100 | EPR685 | ab133616 | Cyan (480) |
| 3 | **CD8** | Leica | RTU | 4B11 | PA0183 | Yellow (520) |
| 4 | **CD68** | Biogenex | RTU | KP1 | AM416-5M | Magenta (690) |
| 5 | **HLA-DR** | Abcam | 1:200 | LN-3 | ab166777 | Green (570) |
| 6 | **FOXP3** | Abcam | 1:200 | 236A/E7 | ab20034 | Red (620) |
| 7 | **NKX3.1** | Cell Marque | 1:50 | EP356 | 441R-15 | White (780) |

**Supplemental Table S2.** Determined cell types:

| Cell | Marker |
| --- | --- |
| Epithelial cells (Tumor) | NKX3.1+ |
| Stromal cells | NKX3.1- |
| CTLs | CD8+CD4- |
| T-helper cells | CD4+CD8-FOXP3- |
| CD4 Tregs | CD4+CD8-FOXP3+ |
| Macrophages | CD68+ |
| M1 Macrophages | CD68+HLA-DR+ |
| M2- Macrophages | CD68+HLA-DR- |

**Supplemental Table S3.** Spatial distribution: Distances determined:

| Distance | Marker |
| --- | --- |
| CTLs to tumor | CD8+CD4- to NKX3.1+ |
| CTLs to CD68+ macrophages | CD8+CD4- to CD68+ |
| T-helper/CTL dyads to tumor | CD8+CD4- and CD4+FOXP3- to NKX3.1 |
| CTLs to M2 macrophages | CD8+CD4- to CD68+HLA-DR- |
| M1 macrophages to tumor | CD68+HLA-DR+ to NKX3.1+ |
| M2 macrophages to tumor | CD68+HLA-DR- to NKX3.1+ |
| CD4 Tregs to tumor | CD4+CD8-FOXP3+ to NKX3.1+ |

**Supplemental Table S4.** Qualitative assessment of staining intensity of all ROIs for each resection specimen of nADT groups.

| Phenotypes | Staining intensity | ≤ 2 month ADT (n=18) | 3-5 month ADT (n=14) | ≥ 6 month ADT (n=11) |
| --- | --- | --- | --- | --- |
| **ROIs/specimen** | Mean (SEM) | 28.8 (3.4) | 25.4 (4.6) | 26.5 (5.5) |
| **M2 abundance** | 1+ | 10 | 8 | 3 |
|  | 2+ | 6 | 4 | 4 |
|  | 3+ | 2 | 2 | 5 |
| **CTL abundance** | 1+ | 9 | 3 | 7 |
|  | 2+ | 7 | 5 | 3 |
|  | 3+ | 2 | 6 | 1 |

## Supplemental Figures

**Supplemental Figure S1.**
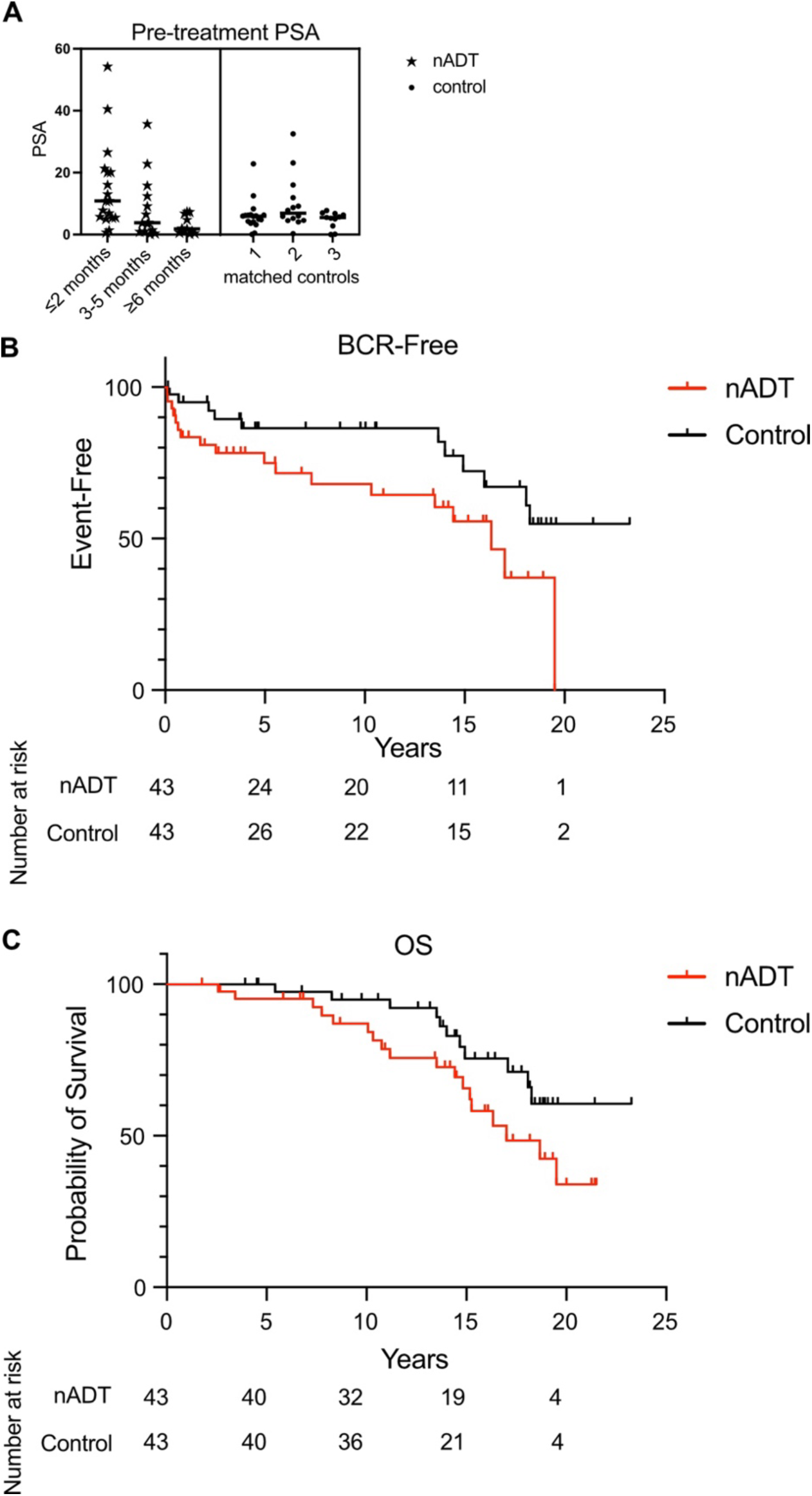
Characteristics and outcomes for the cohorts. **A.** Circulating PSA levels in the three nADT duration groups and their corresponding matched controls prior to therapy. Kruskal-Wallis testing reveals no differences in PSA levels amongst the nADT treated groups, or between nADT and matched control groups. Kaplan Meier curves showing **(B)** Biochemical recurrence-free proportion and **(C)** Overall Survival (OS), with the number at risk in the nADT and control cohorts indicated below. Log-rank testing revealed P = 0.036 for BCR and 0.984 for OS.

**Supplemental Figure S2.**
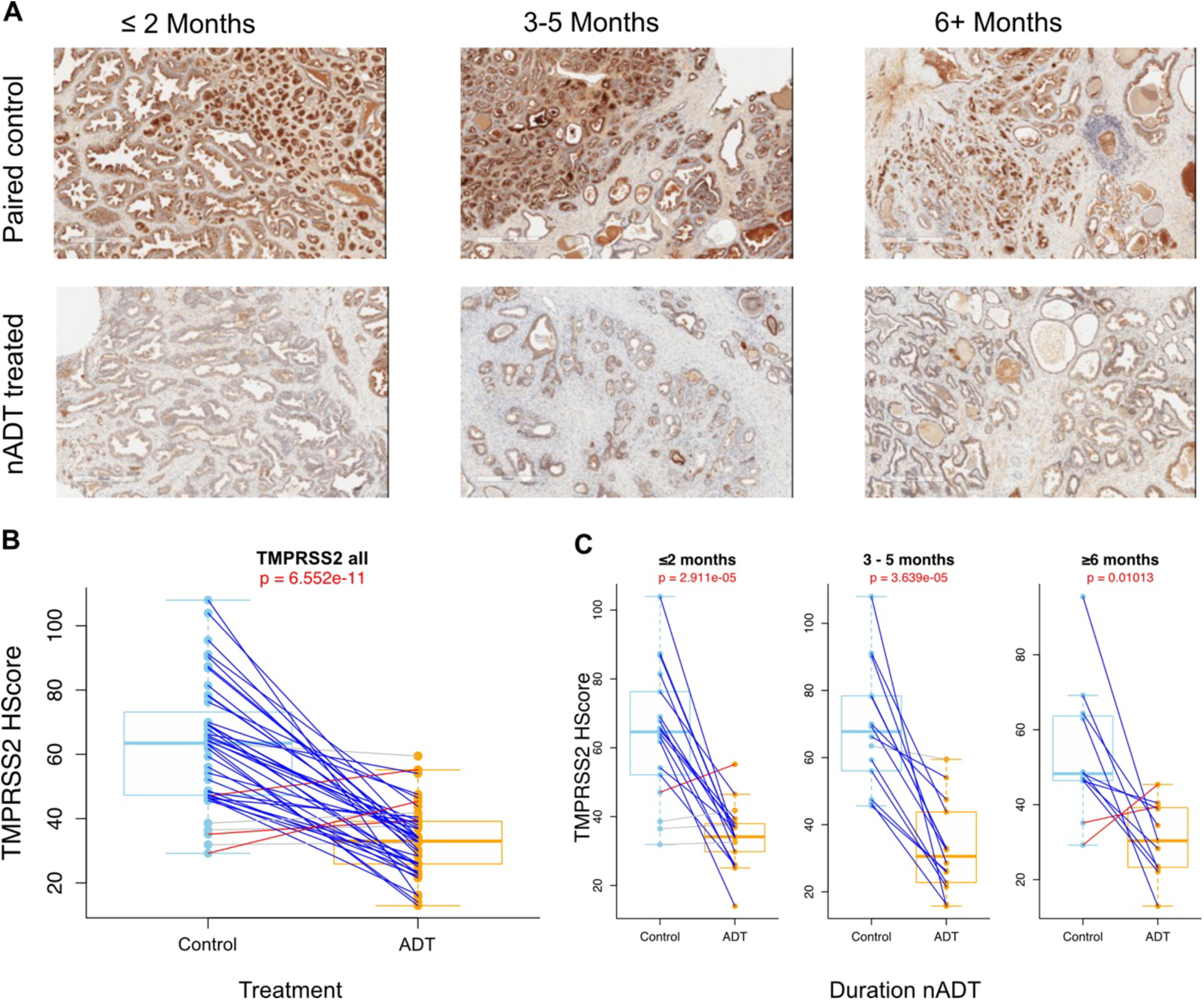
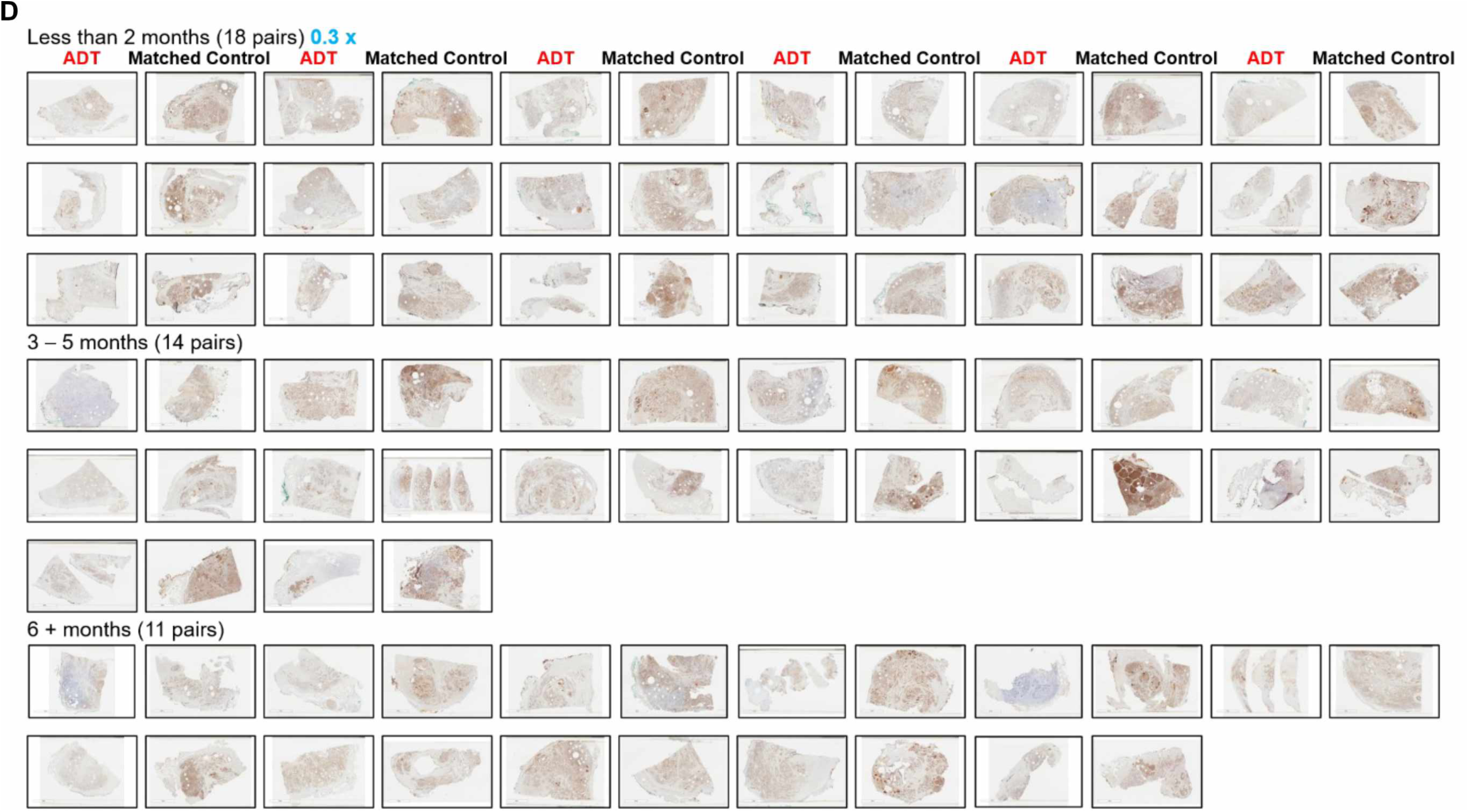
TMPRSS2 protein levels are reduced after nADT. **A.** Representative TMPRSS2 IHC staining (brown) of specimens from each nADT duration group and their corresponding matched controls. **B.** Quantification of TMPRSS2 staining of nADT cohort (orange, n = 43) compared to matched controls (light blue, n = 43). **C.** Quantification of TMPRSS2 staining of the nADT duration groups tissues (orange, n = 18, 14, 11) compared to their matched control tissues (light blue, n = 18, 14, 11). Red lines connecting pairs indicate a > 10% increase in expression, while blue lines indicate a > 10% decrease after nADT (gray lines indicate < 10% change). H score differences determined using paired T-tests in R. **D (below)**. Aperio scanned whole slide images of TMPRSS2 Immunohistochemistry staining of prostate resection specimens from the entire cohort (n = 86). Matched pairs (columns) grouped by duration of nADT (rows).

**Supplemental Figure S3.**
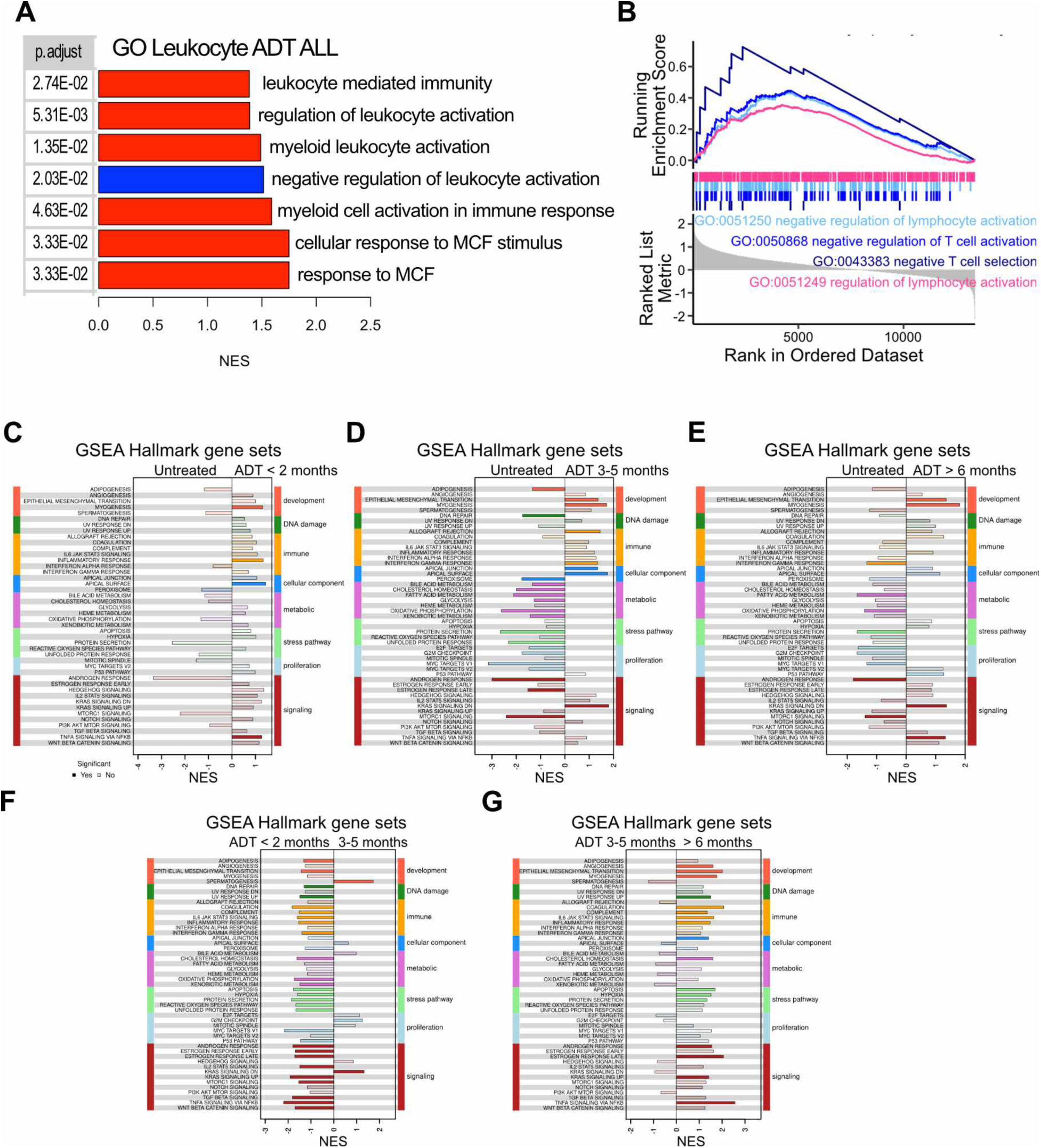
Gene set enrichment analysis by nADT duration. **A.** GSEA of GO Leukocyte pathways for nADT cohort versus matched control cases (43 Pairs). **B.** Enrichment of GO pathways regulating T cell activation nADT cohort versus matched control cases (43 Pairs). **C.** GSEA Hallmark analysis of patients receiving nADT for less than two months compared to paired control (18 pairs). **D.** GSEA Hallmark analysis of patients receiving nADT between 3 to 5 months compared to paired control (14 pairs). **E.** GSEA Hallmark analysis of patients receiving nADT for more than 6 months compared to paired control (11 pairs). The deep color represented significant enrichment while the light color represented the unsignificant change of pathway. **F.** GSEA Hallmark analysis comparing 2 or fewer months versus 3-5 months nADT. **G.** GSEA Hallmark analysis comparing 3-5 months versus 6 or more months nADT.

**Supplemental Figure S4.**
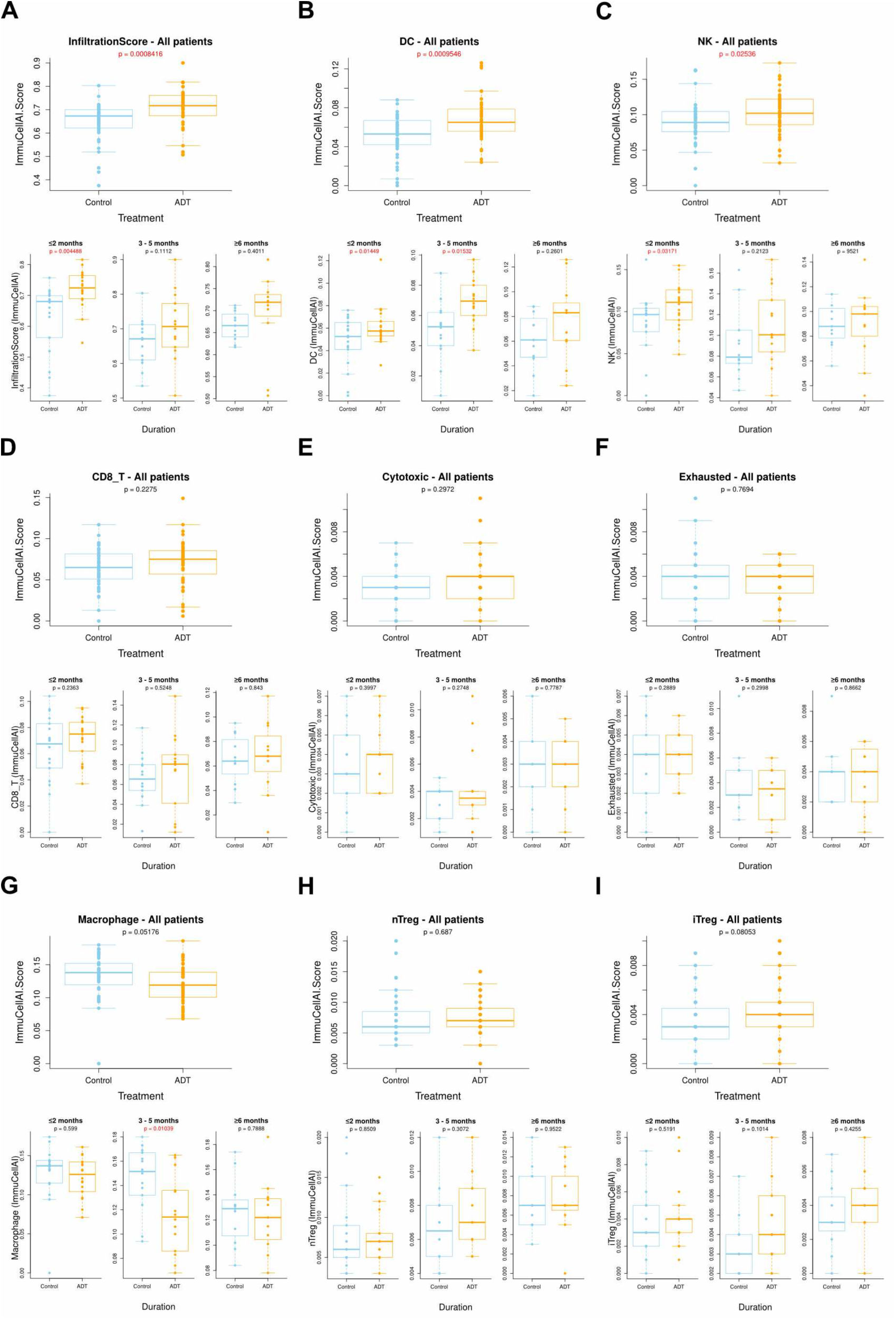
nADT altered immune cell infiltration. Comparison of ImmucelAI imputed immune cell populations for nADT cohort versus matched control cases (43 Pairs, top), and for the three duration of nADT groups: (bottom, ≤ 2 month (18 pairs), 3-5 months (14 pairs), and ≥ 6 month (11 pairs))**. A.** InfiltrationScore. **B.** DC cells. **C.** NK cells. **D.** CD8^+^ T cells. **E.** Cytotoxic T cells. **F.** Exhausted T cells. **G.** Macrophages. **H.** nTregs. **I.** iTregs.

**Supplemental Figure S5.**
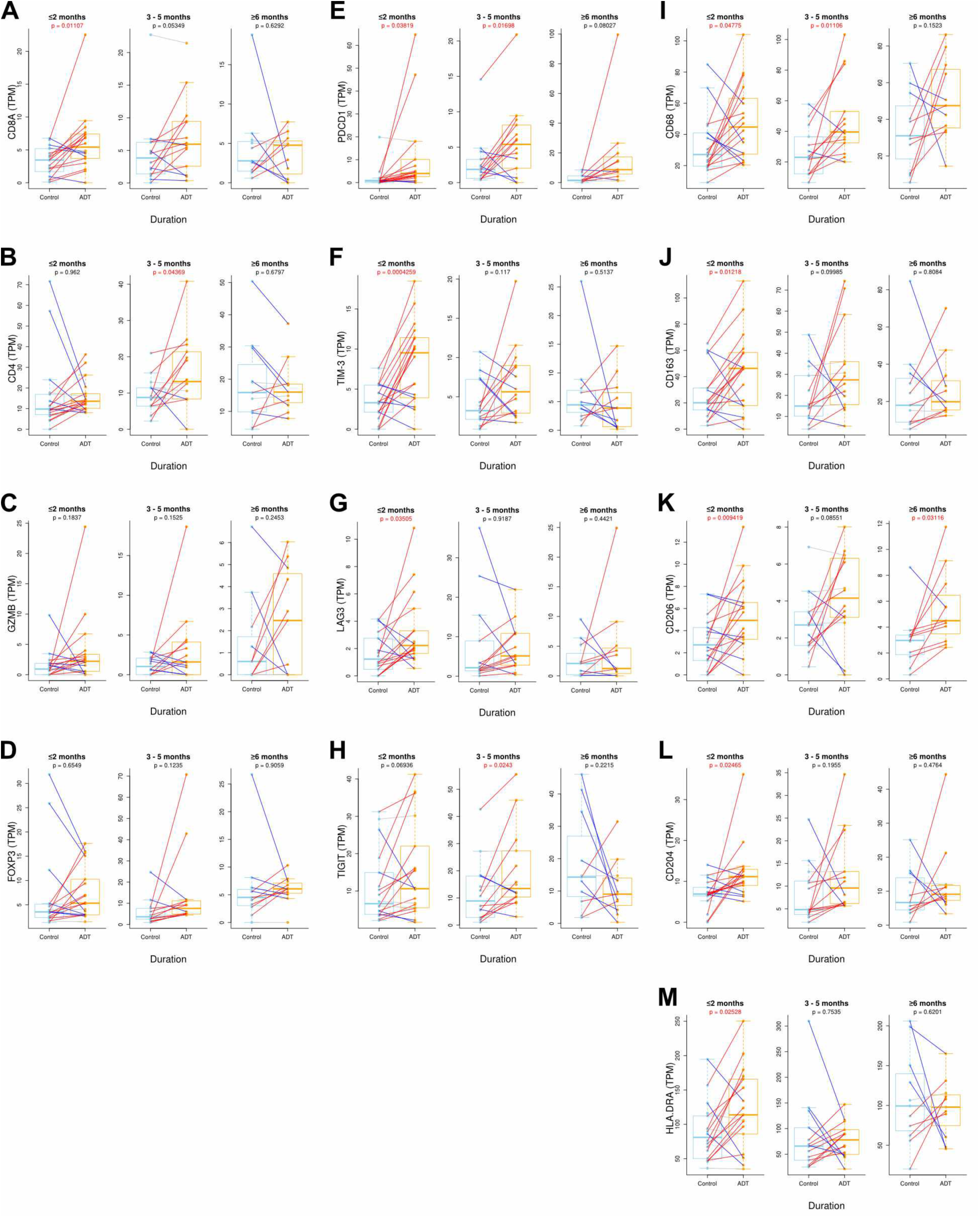
Immune marker gene expression after nADT. Comparison of transcripts per million (TPM) of the indicated genes in paired nADT (orange) and matched control tissues (blue) on for the ≤ 2 months (18 pairs), 3-5 months (14 pairs), and ≥6 months (11 pairs) duration of nADT. Red lines connecting pairs indicate a > 10% increase in expression, while blue lines indicate a > 10% decrease after nADT. Paired t-test p-values are marked in red if <0.05.

**Supplemental Figure S6.**
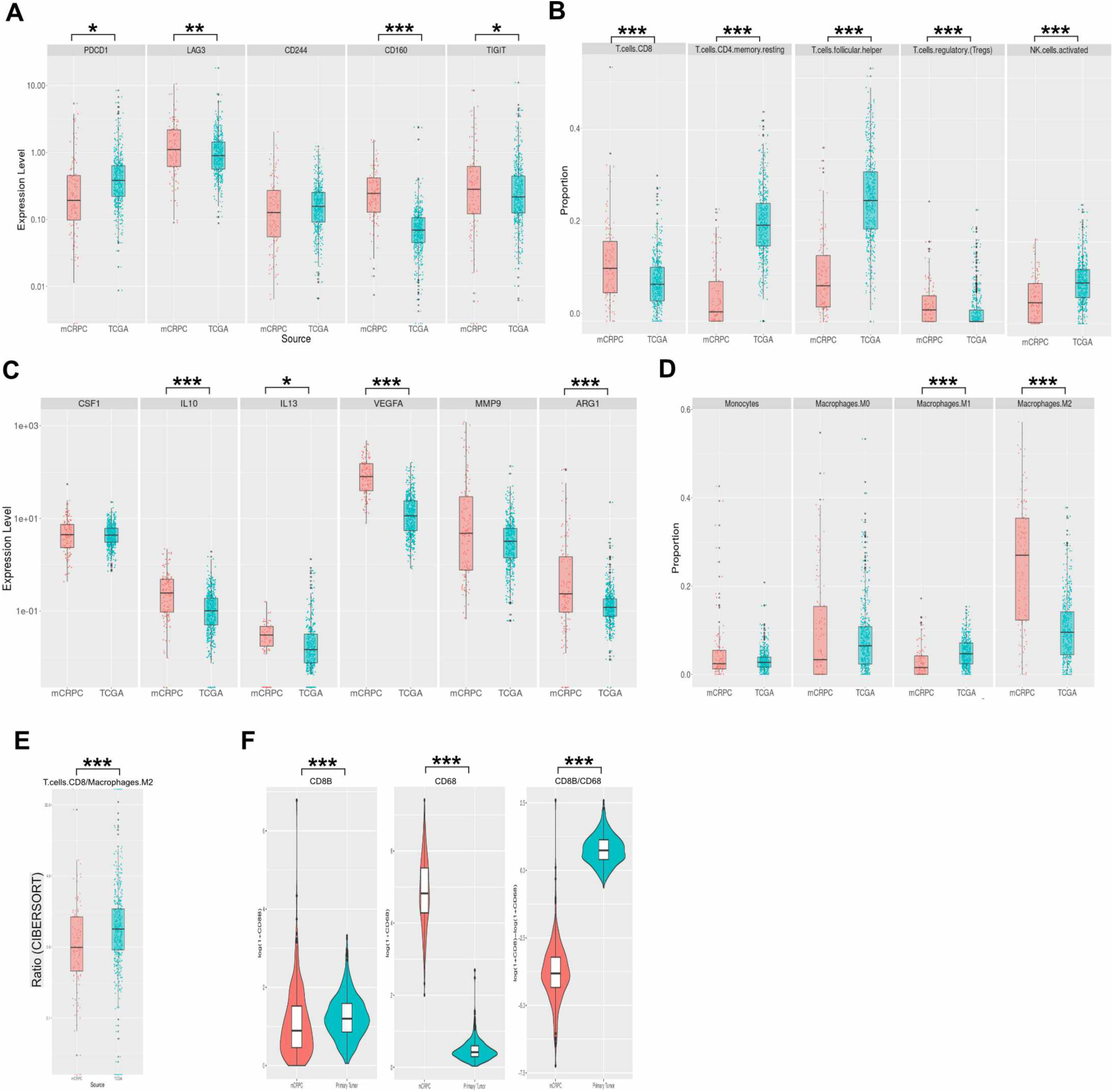
TiME marker profiles shifts to immunosuppressive during progression from localized PrCa to CRPC. Immune cell marker gene relative mRNA levels (**A, C, F**) and estimated immune cell proportion using CIBERSORT analysis (**B, D, E**) of public datasets of castration resistant prostate cancer (CRPC, red) and early localized PrCa (TCGA, green). **A.** Expression of checkpoint genes PDCD1, LAG3, CD244, CD160, TIGIT. **B.** Estimated cell proportions of CD8 T cells, CD4 T memory, T helper cells, Tregs, and activated NK cells. **C.** Expression of CSF1, IL-10, IL-13, VEGFA, MMP99, ARG1 genes. **D.** CIBERSORT cell proportions of monocytes, and M0, M1, and M2 macrophages. **E.** Ratio of CIBERSORT cell proportions of CD8 T cells relative to M2 macrophages. **F.** Expression of CD8B and CD68 genes, and the ratio of CD8B/CD68 gene expression.

**Supplemental Figure S7.**
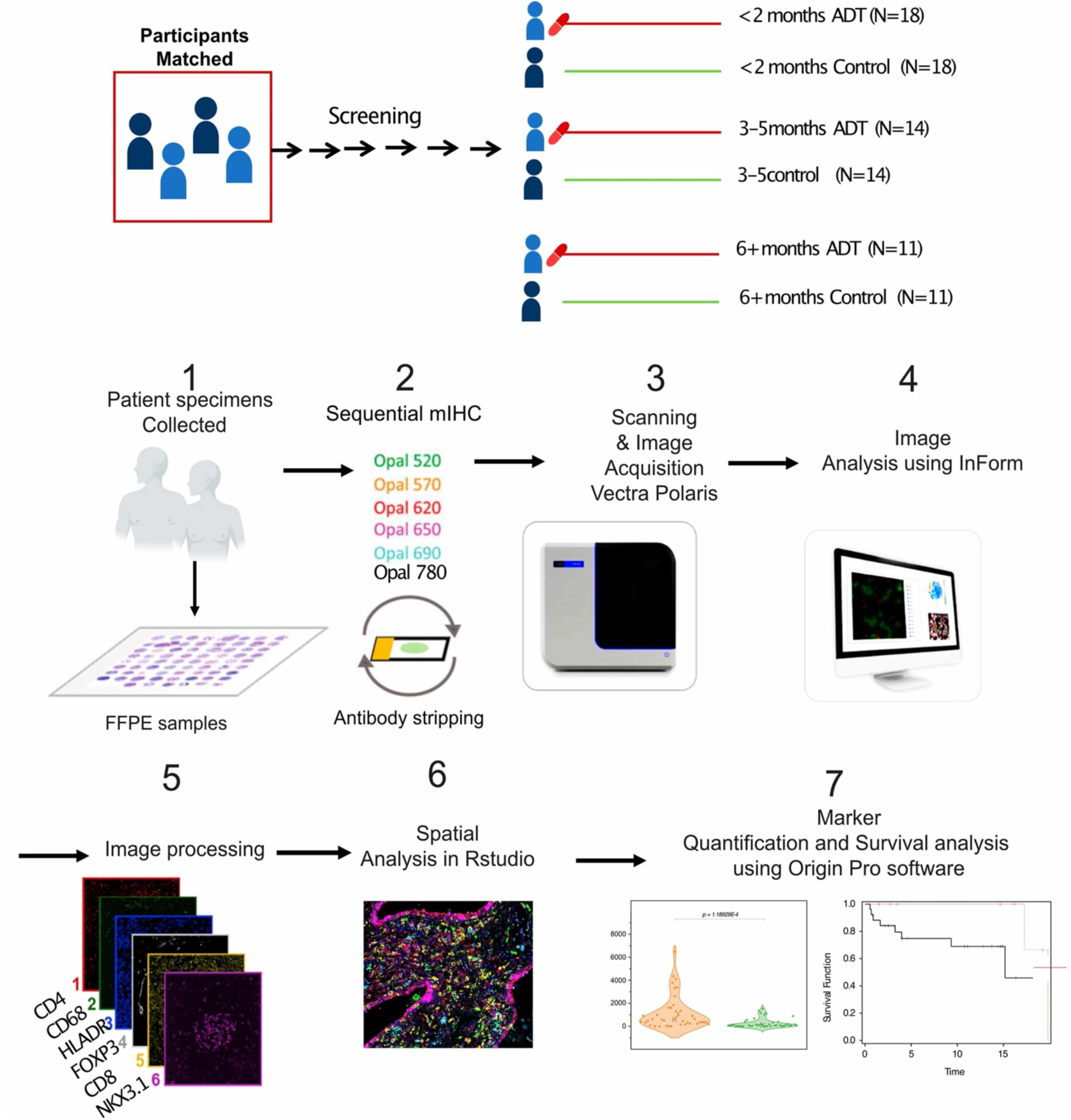
Diagram of cohorts and specimen qmIF analysis. **Top.** Matched cases were separated into groups based on duration of nADT. **1.** A preliminary quality control was done by a pathologist to optimize the preparation of tissue for multiplex immunofluorescence staining and ultimately to guaranty a quality data. **2.** Multiplex IF analysis with tyramide signal amplification was performed on patient samples. The mIF panel (Supplemental Table S1) was stained with the automated Vectra platform. **3.** After staining, slides were imaged using the Vectra 3.0 automated imaging system. **4.** 1744 individual 1 mm^2^ ROIs encompassing the entirety of the 86 specimens were analyzed to better represent the whole tumor, including stroma. **5.** Automated detection and segmentation of specific tissue compartments was performed using InForm software. **6.** The cellular neighborhood identification was performed using phenoptrReport. The distance between each cell and its nearest cell of both the same cell phenotype and the other cell phenotype were automatically calculated and the mean was determined. 7. Phenotyping data produced by inForm machine learning software with spatial map viewer, and differences determined using Wilcoxon Rank-Sum Testing

**Supplemental Figure S8.**
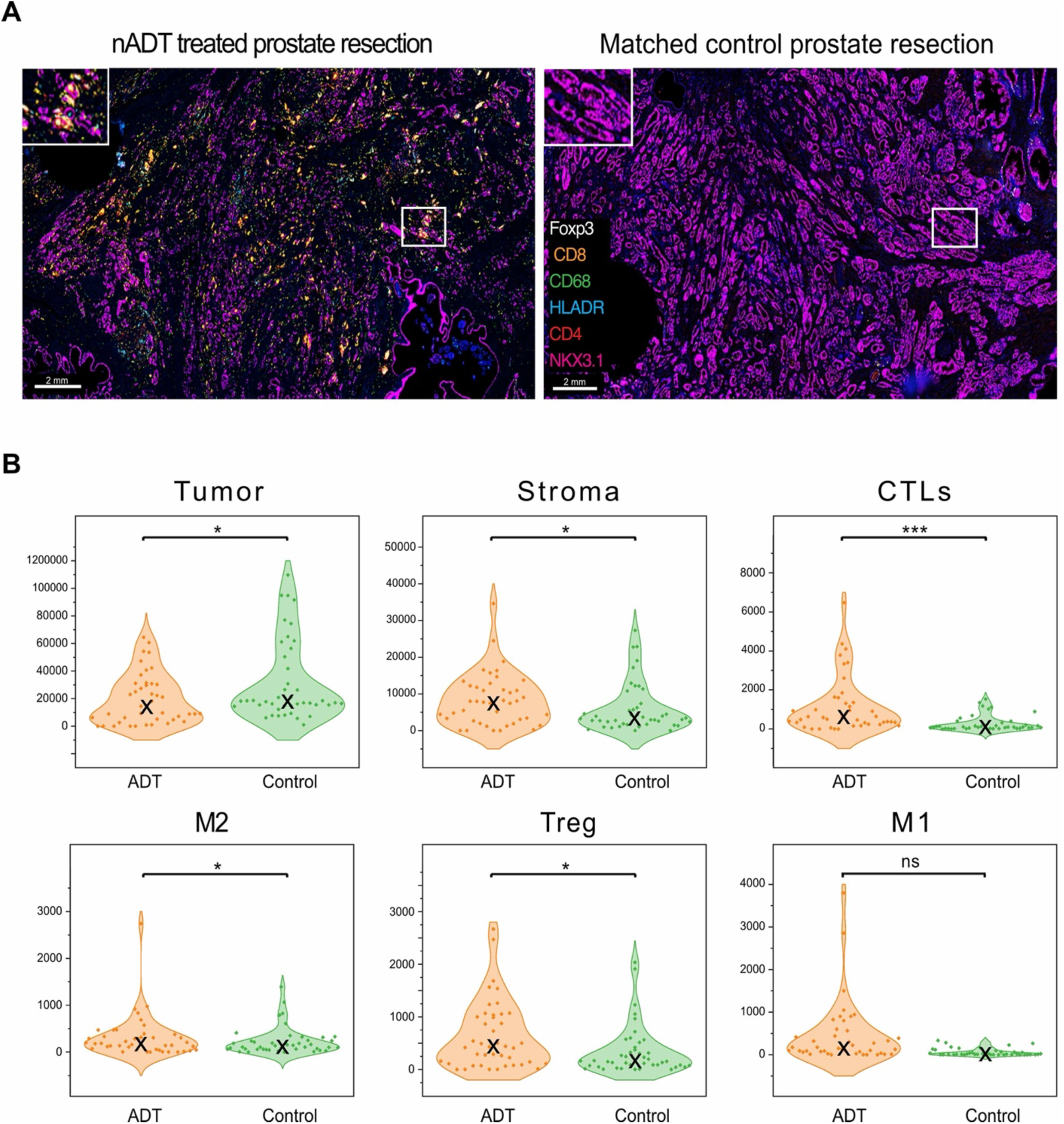
Quantity of tumor, stromal, and immune cells in PrCa are altered by nADT. **A.** Representative low-power fields of prostate tumor resection specimens from patients treated with nADT and matched control, stained for DAPI^+^ cells and tumor (NKX3.1^+^, pink); stroma (marker negative); CTLs (CD8^+^, orange); M2-like TAMs (CD68^+^, HLA-DR^-^, green); Tregs (CD4^+^, Foxp3^+^, white); M1-like TAMs (CD68^+^, HLA-DR^+^, cyan). Marker identified by color indicated in inset, scale bar represents 2 mm. **B.** Cell density (cells/mm^2^) in nADT (n=43, 1077 ROIs quantitated) and control cohorts Vectra (n=43, 667 ROIs quantitated) of tumor, stroma, CTLs, M2, Treg, M1. Means indicated by “X” in violin plot, and comparisons made using the Wilcoxon signed-ranks test. * indicates p-value < 0.05; ** indicates p-value < 0.01; *** indicates p-value < 0.001, ns indicates p> 0.1.

**Supplemental Figure S9.**
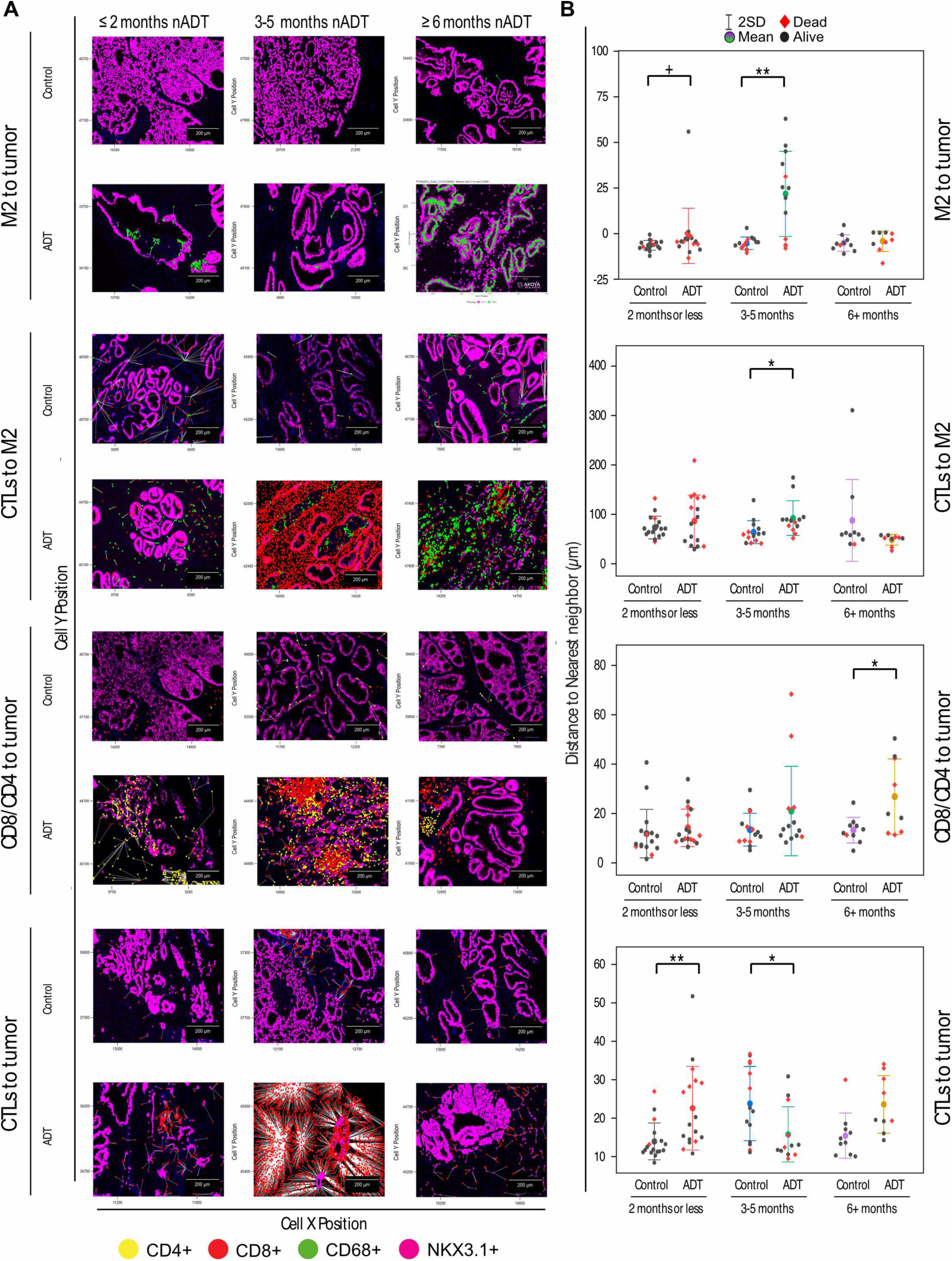
Longer duration nADT increased M2 proximity and reduced CTL proximity to tumor cells. **A**. Example InForm cell spatial mapping images nearest to the mean proximities mapped (white lines) for the indicated cell interaction pairs from patient tissues of ≤ 2 month, 3-5 month, ≥ 6 month nADT groups and matched control tissues. **B.** Graphs of the mean proximities (larger circles, whiskers are SD) of the indicated cell populations in tissues from each duration group and their match controls. Individual patients indicated by small circles. Patients not alive at follow-up is indicated by red colored circles. Specimens/ROIs quantitated: ≤2 month nADT = 18/429; matched controls = 18/322; 3-5 month nADT = 14/356 ROIs; matched controls = 14/187; ≥ 6 month nADT = 11/292; matched controls = 11/158. Each small circles represents the mean proximity derived from an individual patient specimen, with comparisons made using the Wilcoxon signed-ranks test (^+^ indicates p-value < 0.1, * indicates p-value < 0.05; ** indicates p-value < 0.01).

**Supplemental Figure S10.**
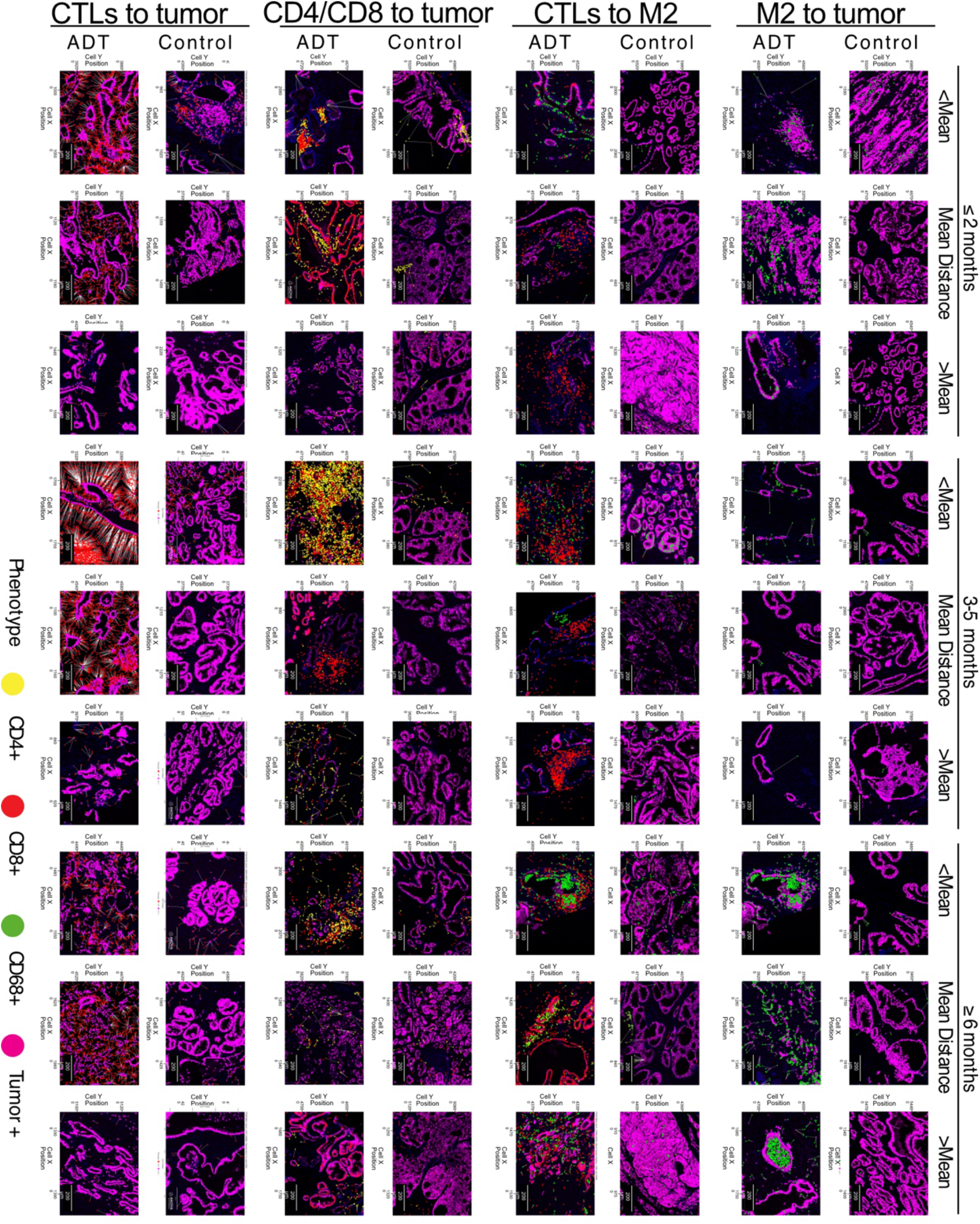
Longer duration nADT increased M2 proximity and reduced CTL proximity to tumor cells. ROIs from three additional in resection specimens from each group illustrating the diversity of spatial proximities between the indicated cell types. Columns are pairs of ROIs from patient tissues at ≤ 2 months, 3-5 months, ≥ 6 months nADT and matched control tissues. Rows show images with proximity measures 1SD above or below the mean (<Mean, >Mean), and an additional example from the case second nearest the mean (Mean Distance). Examples nearest the mean are displayed in Supplemental Fig. S9, and Fig. 5. Qualitative assessment of staining intensity of all ROIs for each resection specimen of nADT groups is detailed in Supplemental Table S4.

**Supplemental Figure S11.**
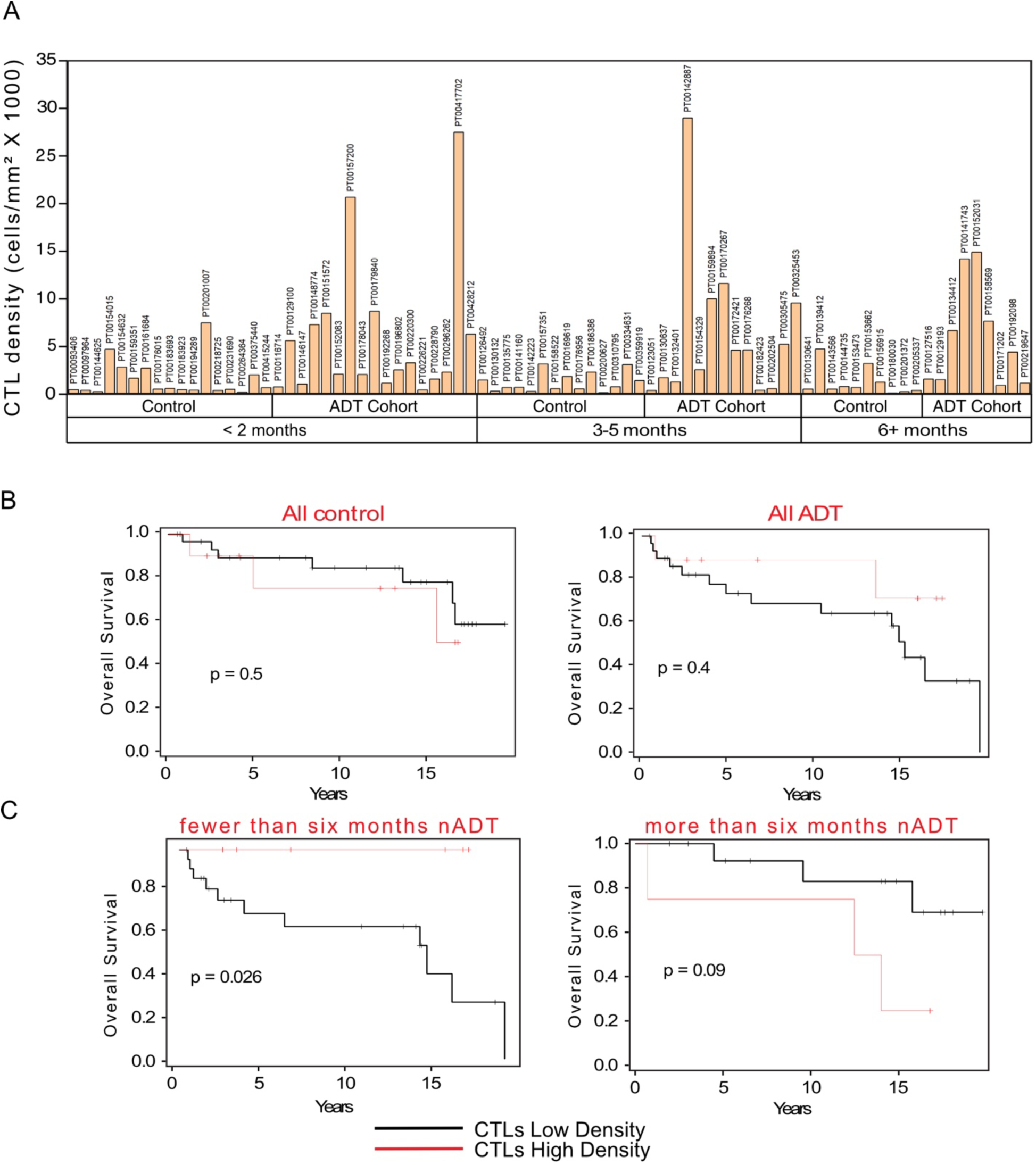
High CTL density was prognostic for overall survival in short duration nADT groups. A. CTL density for each subject. **B.** Kaplan-Meyer curves showing overall survival for the low and high CTL density sub-groups of the control and nADT cohort. **C.** OS for the low and high CTL density groups in patients treated with fewer or more than six months nADT. High CTL density is the upper 25^th^ percentile, Low CTL density is the lower 75^th^ percentile.

**Supplemental Figure S12.**
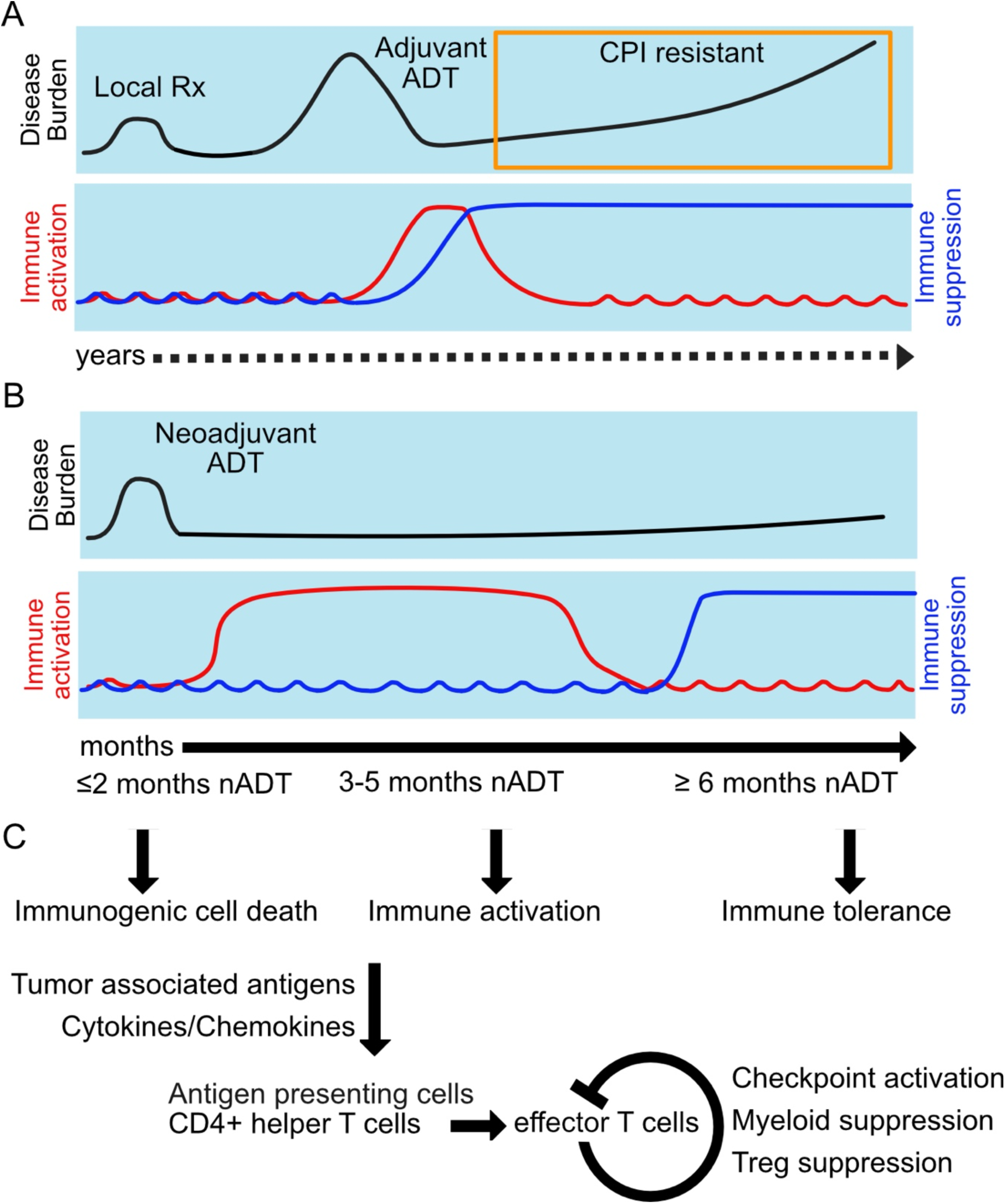
Tumor immune microenvironment response to ADT. **A.** Top: Clinical response (disease burden) of recurrent prostate cancer to adjuvant ADT and later checkpoint inhibitor (CPI) therapy. Bottom: Immune response to adjuvant ADT and approximate timeline. **B.** Top: Clinical response of localized prostate cancer to neoadjuvant ADT (nADT). Bottom: Immune response to nADT and approximate timeline. For both adjuvant ADT and nADT, the effect of immune activation (red line) is transient as the immune activation drives multiple mechanisms of immune suppression (blue line). **C.** Immune changes in response of human prostate tumors to nADT. Levels of tumor-associated antigens and immune regulating cytokines and chemokines are increased by immunogenic cell death, leading to immune activation via triads of antigen-presenting cells and CD4+ T helper cells to induce CD8+ cytotoxic T cells 3-5 months after nADT. Later immune tolerance is characterized by T-cell checkpoint protein expression, myeloid-derived suppressor cells (MDSCs, TAMs) and immunosuppressive Tregs.

## References

1. Antonarakis ES, Piulats JM, Gross-Goupil M, Goh J, Ojamaa K, Hoimes CJ, et al. Pembrolizumab for Treatment-Refractory Metastatic Castration-Resistant Prostate Cancer: Multicohort, Open-Label Phase II KEYNOTE-199 Study. J Clin Oncol 2020;38:395–405.

2. Powles T, Yuen KC, Gillessen S, Kadel EE, 3rd, Rathkopf D, Matsubara N, et al. Atezolizumab with enzalutamide versus enzalutamide alone in metastatic castration-resistant prostate cancer: a randomized phase 3 trial. Nat Med 2022;28:144–53.

3. Sharma P, Pachynski RK, Narayan V, Flechon A, Gravis G, Galsky MD, et al. Nivolumab Plus Ipilimumab for Metastatic Castration-Resistant Prostate Cancer: Preliminary Analysis of Patients in the CheckMate 650 Trial. Cancer Cell 2020;38:489–99 e3.

4. Cristescu R, Mogg R, Ayers M, Albright A, Murphy E, Yearley J, et al. Pan-tumor genomic biomarkers for PD-1 checkpoint blockade-based immunotherapy. Science 2018;362

5. Chen DS, Mellman I. Elements of cancer immunity and the cancer-immune set point. Nature 2017;541:321–30.

6. Yang Y, Attwood K, Bshara W, Mohler JL, Guru K, Xu B, et al. High intratumoral CD8(+) T-cell infiltration is associated with improved survival in prostate cancer patients undergoing radical prostatectomy. Prostate 2021;81:20–8.

7. Espinosa-Carrasco G, Chiu E, Scrivo A, Zumbo P, Dave A, Betel D, et al. Intratumoral immune triads are required for immunotherapy-mediated elimination of solid tumors. Cancer Cell 2024;42:1202–16 e8.

8. Hoffmann JP, Liu JA, Seddu K, Klein SL. Sex hormone signaling and regulation of immune function. Immunity 2023;56:2472–91.

9. Yanai Y, Kosaka T, Mikami S, Arai M, Watanabe K, Takeda T, et al. Dynamics in the Prostate Immune Microenvironment Induced by Androgen Deprivation Therapy. Prostate 2025;85:308–14.

10. Mercader M, Bodner BK, Moser MT, Kwon PS, Park ES, Manecke RG, et al. T cell infiltration of the prostate induced by androgen withdrawal in patients with prostate cancer. Proc Natl Acad Sci U S A 2001;98:14565–70.

11. Gannon PO, Poisson AO, Delvoye N, Lapointe R, Mes-Masson AM, Saad F. Characterization of the intra-prostatic immune cell infiltration in androgen-deprived prostate cancer patients. Journal of immunological methods 2009;348:9–17.

12. Chesner LN, Polesso F, Graff JN, Hawley JE, Smith AK, Lundberg A, et al. Androgen Receptor Inhibition Increases MHC Class I Expression and Improves Immune Response in Prostate Cancer. Cancer Discov 2025;15:481–94.

13. Nesslinger NJ, Sahota RA, Stone B, Johnson K, Chima N, King C, et al. Standard treatments induce antigen-specific immune responses in prostate cancer. Clin Cancer Res 2007;13:1493–502.

14. Morse MD, McNeel DG. Prostate cancer patients on androgen deprivation therapy develop persistent changes in adaptive immune responses. Hum Immunol 2010;71:496–504.

15. Long X, Hou H, Wang X, Liu S, Diao T, Lai S, et al. Immune signature driven by ADT-induced immune microenvironment remodeling in prostate cancer is correlated with recurrence-free survival and immune infiltration. Cell death & disease 2020;11:779.

16. Obradovic AZ, Dallos MC, Zahurak ML, Partin AW, Schaeffer EM, Ross AE, et al. T-Cell Infiltration and Adaptive Treg Resistance in Response to Androgen Deprivation With or Without Vaccination in Localized Prostate Cancer. Clin Cancer Res 2020;26:3182–92.

17. Erlandsson A, Lundholm M, Watz J, Bergh A, Petrova E, Alamdari F, et al. Infiltrating immune cells in prostate cancer tissue after androgen deprivation and radiotherapy. Int J Immunopathol Pharmacol 2023;37:3946320231158025.

18. Dallos MC, Obradovic AZ, McCann P, Chowdhury N, Pratapa A, Aggen DH, et al. Androgen Deprivation Therapy Drives a Distinct Immune Phenotype in Localized Prostate Cancer. Clin Cancer Res 2024;30:5218–30.

19. Erlandsson A, Carlsson J, Lundholm M, Falt A, Andersson SO, Andren O, et al. M2 macrophages and regulatory T cells in lethal prostate cancer. Prostate 2019;79:363–9.

20. Sharma P, Siddiqui BA, Anandhan S, Yadav SS, Subudhi SK, Gao J, et al. The Next Decade of Immune Checkpoint Therapy. Cancer Discov 2021;11:838–57.

21. Dobin A, Davis CA, Schlesinger F, Drenkow J, Zaleski C, Jha S, et al. STAR: ultrafast universal RNA-seq aligner. Bioinformatics 2013;29:15–21.

22. Li B, Dewey CN. RSEM: accurate transcript quantification from RNA-Seq data with or without a reference genome. BMC Bioinformatics 2011;12:323.

23. Robinson MD, McCarthy DJ, Smyth GK. edgeR: a Bioconductor package for differential expression analysis of digital gene expression data. Bioinformatics 2010;26:139–40.

24. Ritchie ME, Phipson B, Wu D, Hu Y, Law CW, Shi W, et al. limma powers differential expression analyses for RNA-sequencing and microarray studies. Nucleic Acids Res 2015;43:e47.

25. Newman AM, Liu CL, Green MR, Gentles AJ, Feng W, Xu Y, et al. Robust enumeration of cell subsets from tissue expression profiles. Nat Methods 2015;12:453–7.

26. Miao YR, Zhang Q, Lei Q, Luo M, Xie GY, Wang H, et al. ImmuCellAI: A Unique Method for Comprehensive T-Cell Subsets Abundance Prediction and its Application in Cancer Immunotherapy. Adv Sci (Weinh) 2020;7:1902880.

27. Subramanian A, Tamayo P, Mootha VK, Mukherjee S, Ebert BL, Gillette MA, et al. Gene set enrichment analysis: a knowledge-based approach for interpreting genome-wide expression profiles. Proc Natl Acad Sci U S A 2005;102:15545–50.

28. Liberzon A, Birger C, Thorvaldsdottir H, Ghandi M, Mesirov JP, Tamayo P. The Molecular Signatures Database (MSigDB) hallmark gene set collection. Cell Syst 2015;1:417–25.

29. Korotkevich G, Sukhov V, Budin N, Shpak B, Artyomov MN, Sergushichev A. Fast gene set enrichment analysis. bioRxiv 2021

30. Wu T, Hu E, Xu S, Chen M, Guo P, Dai Z, et al. clusterProfiler 4.0: A universal enrichment tool for interpreting omics data. Innovation (Camb) 2021;2:100141.

31. Isse K, Lesniak A, Grama K, Roysam B, Minervini MI, Demetris AJ. Digital transplantation pathology: combining whole slide imaging, multiplex staining and automated image analysis. Am J Transplant 2012;12:27–37.

32. Salles DC, Vidotto T, Faisal FA, Tosoian JJ, Guedes LB, Muranyi A, et al. Assessment of MYC/PTEN Status by Gene-Protein Assay in Grade Group 2 Prostate Biopsies. J Mol Diagn 2021;23:1030–41.

33. Lucas JM, True L, Hawley S, Matsumura M, Morrissey C, Vessella R, et al. The androgen-regulated type II serine protease TMPRSS2 is differentially expressed and mislocalized in prostate adenocarcinoma. J Pathol 2008;215:118–25.

34. Lucas JM, Heinlein C, Kim T, Hernandez SA, Malik MS, True LD, et al. The androgen-regulated protease TMPRSS2 activates a proteolytic cascade involving components of the tumor microenvironment and promotes prostate cancer metastasis. Cancer Discov 2014;4:1310–25.

35. Cancer Genome Atlas Research N. The Molecular Taxonomy of Primary Prostate Cancer. Cell 2015;163:1011–25.

36. Robinson D, Van Allen EM, Wu YM, Schultz N, Lonigro RJ, Mosquera JM, et al. Integrative clinical genomics of advanced prostate cancer. Cell 2015;161:1215–28.

37. Abida W, Cyrta J, Heller G, Prandi D, Armenia J, Coleman I, et al. Genomic correlates of clinical outcome in advanced prostate cancer. Proc Natl Acad Sci U S A 2019;116:11428–36.

38. Shaw GL, Whitaker H, Corcoran M, Dunning MJ, Luxton H, Kay J, et al. The Early Effects of Rapid Androgen Deprivation on Human Prostate Cancer. Eur Urol 2016;70:214–8.

39. Mostaghel EA, Page ST, Lin DW, Fazli L, Coleman IM, True LD, et al. Intraprostatic androgens and androgen-regulated gene expression persist after testosterone suppression: therapeutic implications for castration-resistant prostate cancer. Cancer Res 2007;67:5033–41.

40. Galluzzi L, Buque A, Kepp O, Zitvogel L, Kroemer G. Immunogenic cell death in cancer and infectious disease. Nat Rev Immunol 2017;17:97–111.

41. Rapoport BL, Anderson R. Realizing the Clinical Potential of Immunogenic Cell Death in Cancer Chemotherapy and Radiotherapy. Int J Mol Sci 2019;20

42. Westbrook TC, Guan X, Rodansky E, Flores D, Liu CJ, Udager AM, et al. Transcriptional profiling of matched patient biopsies clarifies molecular determinants of enzalutamide-induced lineage plasticity. Nat Commun 2022;13:5345.

43. Linder S, Hoogstraat M, Stelloo S, Eickhoff N, Schuurman K, de Barros H, et al. Drug-Induced Epigenomic Plasticity Reprograms Circadian Rhythm Regulation to Drive Prostate Cancer toward Androgen Independence. Cancer Discov 2022;12:2074–97.

44. Aras S, Zaidi MR. TAMeless traitors: macrophages in cancer progression and metastasis. Br J Cancer 2017;117:1583–91.

45. Nonomura N, Takayama H, Nakayama M, Nakai Y, Kawashima A, Mukai M, et al. Infiltration of tumour-associated macrophages in prostate biopsy specimens is predictive of disease progression after hormonal therapy for prostate cancer. BJU Int 2011;107:1918–22.

46. Guan X, Polesso F, Wang C, Sehrawat A, Hawkins RM, Murray SE, et al. Androgen receptor activity in T cells limits checkpoint blockade efficacy. Nature 2022;606:791–6.

47. Pelka K, Hofree M, Chen JH, Sarkizova S, Pirl JD, Jorgji V, et al. Spatially organized multicellular immune hubs in human colorectal cancer. Cell 2021;184:4734–52 e20.

48. Drake CG, Doody AD, Mihalyo MA, Huang CT, Kelleher E, Ravi S, et al. Androgen ablation mitigates tolerance to a prostate/prostate cancer-restricted antigen. Cancer Cell 2005;7:239–49.

49. Vicier C, Ravi P, Kwak L, Werner L, Huang Y, Evan C, et al. Association between CD8 and PD-L1 expression and outcomes after radical prostatectomy for localized prostate cancer. Prostate 2021;81:50–7.

50. Lyu A, Fan Z, Clark M, Lea A, Luong D, Setayesh A, et al. Evolution of myeloid-mediated immunotherapy resistance in prostate cancer. Nature 2025;637:1207–17.

51. Siddiqui BA, Chapin BF, Jindal S, Duan F, Basu S, Yadav SS, et al. Immune and pathologic responses in patients with localized prostate cancer who received daratumumab (anti-CD38) or edicotinib (CSF-1R inhibitor). J Immunother Cancer 2023;11

52. Di Mitri D, Mirenda M, Vasilevska J, Calcinotto A, Delaleu N, Revandkar A, et al. Re-education of Tumor-Associated Macrophages by CXCR2 Blockade Drives Senescence and Tumor Inhibition in Advanced Prostate Cancer. Cell Rep 2019;28:2156–68 e5.

